# Plasma DNA fragmentomic analysis for evaluation of patients with pancreatic cystic neoplasms

**DOI:** 10.1101/2024.12.13.24318579

**Authors:** Clayton T. Marcinak, Michelle D. Stephens, Jiarui Chen, Nabeel Merali, Bradon R. McDonald, Elise C. Dietmann, Muhammad Talha Nawaz, Stephanie M. McGregor, Sean M. Ronnekleiv-Kelly, Sharon M. Weber, Everlyne N. Nkadori, Peter G. Stadmeyer, Mark E. Benson, Deepak V. Gopal, Syed Nabeel Zafar, Kaitlyn J. Kelly, Jomol Mathew, Ali Pirasteh, Adam E. Frampton, Patrick R. Pfau, Shivan Sivakumar, Rebecca M. Minter, Muhammed Murtaza

## Abstract

**Background:** Incidental detection of pancreatic cystic neoplasms (PCNs) is common, but to identify patients with malignant PCNs (high-grade dysplasia or carcinoma) who need invasive testing such as endoscopic ultrasound (EUS) or surgery is challenging. Plasma DNA fragmentomic analysis has recently shown promise for early detection of pancreatic ductal adenocarcinoma (PDAC).

**Objective:** To evaluate the ability of plasma DNA fragmentomic analysis to differentiate between patients with benign and malignant PCNs.

**Design:** We performed whole genome sequencing of pre-treatment plasma DNA from 121 patients with PDAC, 121 age and sex-matched healthy controls, and 99 patients with PCNs referred for EUS (total n=341). After training and validating a plasma DNA fragmentomics machine learning model for PDAC detection, we independently evaluated its performance for detection of malignant PCNs, and compared with circulating glycoproteins.

**Results:** Eleven of 99 patients (11%) were diagnosed with malignant PCNs during 24.6 months of clinical follow-up. For PDAC detection, plasma DNA analysis showed area under the receiver operating characteristic curve (AUROC) of 0.97. For malignant PCN detection, plasma DNA analysis showed AUROC of 0.67, while CA19-9, CA125 and CEA showed AUROCs of 0.78, 0.54, and 0.63, respectively. Combined analysis of plasma DNA with glycoproteins showed AUROC of 0.92, demonstrating 91% sensitivity, 99% negative predictive value, 88% specificity and 48% positive predictive value for malignant PCN detection.

**Conclusion:** Plasma DNA fragmentomics and glycoprotein biomarkers showed complementary performance for detecting malignant PCNs in patients referred for EUS. This approach could improve patient selection for EUS, and warrants further evaluation.

## Introduction

Pancreatic ductal adenocarcinoma (PDAC) is the third leading cause of cancer death in the United States, with over 50,000 deaths attributed to the disease annually.^1^ While patients diagnosed at earlier stages have better prognosis, there are no cancer screening guidelines for PDAC in the general population due to its low incidence.^2^ However, institutional screening programs for patients at high risk for pancreatic cancer significantly improve disease detection at early stage with reported five-year survival rates of over 70%.^3^ Patients at high risk include those with chronic pancreatitis, a strong family history of pancreatic cancer, new-onset diabetes, and incidentally detected mucinous pancreatic cystic neoplasms (PCNs).

As imaging modalities improve and the population ages, incidental detection of PCNs is increasing. PCNs are detected in an estimated 16% of the general population undergoing imaging, increasing from 9% of individuals less than 60 years of age to 26% of individuals 70 years and older.^4^ Despite their prevalence, the risk of malignant progression is small. One population-based study of PCNs estimated the annual progression rate at 0.47%, with a seven-year cumulative rate of progression to cancer of 3.0%.^5^ In current practice, patients with PCNs undergo longitudinal clinical follow-up, and those with high-risk or worrisome features on magnetic resonance imaging (MRI) such as larger size, thickened cyst walls or mural nodules are evaluated with endoscopic ultrasound (EUS) and fine needle aspiration (FNA) with biopsy when indicated.^6–8^ However, current clinical protocols have limited accuracy for detection of malignant PCNs.^9^ Each year, an estimated 41,000 to 90,000 EUS procedures are performed in the US to evaluate PCNs, and more than 90% of patients are not diagnosed with malignant PCNs (high-grade dysplasia or adenocarcinoma)^10–13^. EUS and FNA require sedation, and are associated with a complication rate of up to 2% including acute pancreatitis, hemorrhage and perforation^14^. Amongst patients who undergo resection following EUS and multidisciplinary evaluation, only 23% of patients are reported to have high-grade dysplasia or invasive cancer.^15^ Hence, there is unmet clinical need to improve selection of patients at the highest risk of malignant PCNs who are most likely to benefit from EUS and surgical resection. In addition, EUS is a critical resource for patients with cancer awaiting tissue diagnosis and decreasing the use of this important modality for patients with benign lesions would improve capacity for patients with more urgent conditions. Thus, improved risk stratification can help reduce unnecessary diagnostic procedures and complex surgeries in patients with benign PCNs, while also reducing financial burden on the healthcare system.^16, 17^

The current role of blood tests for cancer detection and risk stratification in patients with PCNs is limited. Investigation of serum CA19-9 has previously shown low accuracy for detection of malignant PCNs.^18, 19^ Recent studies have demonstrated the promise of circulating tumor DNA analysis for multi-cancer early detection^20^. These approaches rely on identification of somatic mutations^21^, differences in methylation signatures^22^, or differences in plasma DNA fragmentation patterns.^23^ Multiple groups have shown the potential of circulating tumor DNA analysis for distinguishing patients with PDAC from healthy individuals.^24–27^ More recently, analysis of plasma DNA fragmentation patterns has shown promising results for PDAC detection in case-control and prospective studies^28–30^. However, the relevance and performance of these approaches to distinguish benign from malignant lesions in a clinical population of patients with PCNs is unknown.

In the current study, we address this gap and evaluate the performance of plasma DNA fragmentation analysis to distinguish malignant from benign PCNs in a prospective cohort of previously asymptomatic patients with incidentally detected PCNs who were referred for endoscopic evaluation. We first train and cross-validate PDAC-specific machine learning models based on multiple plasma DNA fragmentation features extracted from whole genome sequencing of blood samples in a cohort of patients with PDAC, and age- and sex-matched health controls. We then evaluate the performance of these models independently in patients with PCNs and compare with circulating glycoproteins including CA19-9, CA125 and CEA, for detection of malignancy (high-grade dysplasia or cancer).

## Methods

### Ethics Statement

Samples from patients with PDAC, PCNs and healthy controls were analyzed under ethical approvals by the Institutional Review Board (IRB) of the University of Wisconsin–Madison (protocols 2016-0934, 2022-1665 and 2022-1460) and the NHS Health Research Authority (protocol 20SURN277406). Patients or the public were not involved in the design, or conduct, or reporting, or dissemination plans of our research.

### Training and validating plasma DNA fragmentation analysis for PDAC detection

We and others have recently demonstrated that distribution of plasma DNA fragments across the genome is non-random, and represents chromatin accessibility within the cells where such DNA is shed from^31^. There are multiple features that capture differences in fragmentation patterns between patients with cancer and healthy individuals, and recent publications have described how these features can be leveraged to identify patients with cancer. Here, we evaluate four different sets of fragmentomic features including genome-wide variation in short-to-long fragment ratios^23^, nucleotide motifs at plasma DNA fragment ends^32^, genome-wide variation in normalized sequencing coverage^30^, and an approach that we developed recently called genome-wide analysis of aberrant fragment ends (GALYFRE)^31^. GALYFRE measures DNA fragmentation at unexpected genomic loci to determine if a blood sample was collected from a patient with cancer, and incorporates fraction of plasma DNA fragments with unexpected breakpoints, as well as nucleotide frequencies at fragment ends. In addition to evaluating each feature set independently, we also evaluate a random forest model that combined all features into a single model, allowing the choice of the most informative features from across all four sets.

In the current study, we extracted all four feature sets and trained independent random forest machine models for detection of pancreatic cancer using a cohort of patients diagnosed with PDAC, and age- and sex-matched healthy individuals. For model training and cross-validation, we analyzed pre-treatment plasma samples collected from 121 patients with PDAC (28 patients with stage I, 42 patients with stage II, 30 patients with stage III, and 21 patients with stage IV disease) from the biospecimen repositories of the University of Wisconsin Carbone Cancer Center and the Royal Surrey Hospital NHS Trust, and an equal number of age and sex matched healthy control samples from a community-based longitudinal cohort in Wisconsin^33^. Control samples were confirmed to be cancer-free prior to and since sample collection by linking the survey records with the Wisconsin Cancer Reporting System, the state cancer registry.

### Patients with pancreatic cystic neoplasms

To evaluate biomarker performance in patients with PCNs, we prospectively collected peripheral blood from consecutive patients referred to our institution for endoscopic evaluation of a pancreas lesion incidentally identified on imaging, from 2022 to 2024. Eligible patients were defined as those with at least one of the following characteristics on imaging: (1) a pancreas lesion greater than or equal to 2 cm in greatest dimension with at least a partial cystic component, (2) a dilated main pancreatic duct, defined as a diameter greater than or equal to 5 mm, (3) a change in pancreatic duct diameter, and (4) other high-risk stigmata or worrisome features defined by the Kyoto guidelines.^34^ Patients with a discrete solid mass on imaging and those with signs and symptoms of biliary obstruction were not eligible. Patients continued guideline concordant surveillance and management of their pancreatic lesions through the multidisciplinary UW Health Pancreatic Cancer Prevention Clinic (PCPC). As part of UW PCPC, symptoms, genetic risk, circulating biomarkers, endoscopic and imaging findings from each patient were reviewed by a committee of surgeons, gastroenterologists, radiologists and geneticists, using surveillance protocols driven by American College of Gastroenterology guidelines.^8^

Of 233 eligible patients referred for EUS during the study period, 136 patients were successfully consented and enrolled in the study (**Figure 1)**. Following informed consent, blood samples were collected and analyzed. At the time of final data analysis, patients were excluded if they met any of the following criteria: (1) a history of cancer within the past five years, other than localized, cutaneous squamous cell or basal cell carcinoma, (2) a history of solid organ or bone marrow transplant, (3) a history of a prior high-risk PCN that was resected, or (4) a known germline oncogenic gene mutation. Patients who were found to have a non-epithelial lesion (*e.g.,* gastrointestinal stromal tumor) during evaluation of their PCN were also excluded. The final dataset was comprised of 99 patients with PCNs.

**Figure 1:**
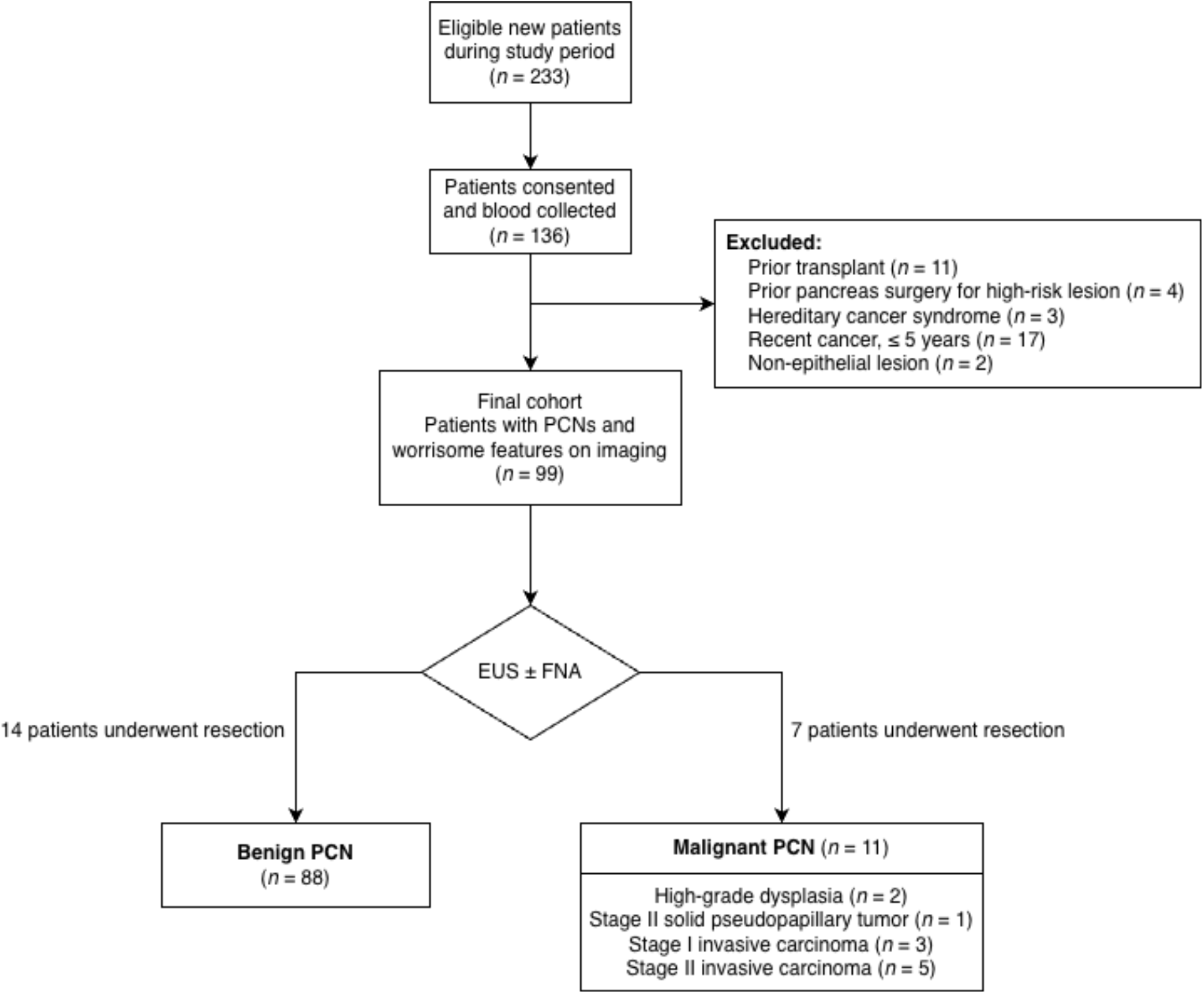
An overview of the study design. A diagram detailing the prospective enrollment, exclusion criteria applied, and number of eligible patients with incidental pancreas lesions. Median time of follow-up for included patients was 24.6 months. Abbreviations: PDAC, pancreatic ductal adenocarcinoma; EUS, endoscopic ultrasound; HGD, high-grade dysplasia.

A patient was considered to have malignant PCN if either high-grade dysplasia or invasive carcinoma were identified on fine needle aspiration, core needle biopsy, or surgical resection specimen. The remaining patients who continued in surveillance were considered to have benign PCN (**Figure 1**). To evaluate the potential impact of clinical false negatives (patients who may be diagnosed with malignancy in the near future) on biomarker performance, we also evaluated all biomarkers in a subgroup of patients with histologically confirmed diagnoses. This subgroup included both patients with a diagnosis of malignant PCN, and those with definitive diagnosis of benign PCN upon surgical resection.

### Analysis of glycoprotein biomarkers and plasma DNA fragmentation

Plasma samples from patients with PCNs were analyzed for three glycoprotein biomarkers previously described to have an association with pancreatic cancer: carbohydrate antigen 19-9 (CA19-9), carcinoembryonic antigen (CEA), and cancer antigen 125 (CA125).^35^ For plasma DNA fragmentation analysis, blood samples were processed to isolate plasma DNA and perform whole genome sequencing (WGS).^31^ Following alignment of sequencing data to the human genome, we extracted four sets of fragmentomic features including 1) fragment lengths and the ratio of short-to-long fragments across the genome, 2) sequencing coverage across the genome, 3) frequency of 4-mer sequence end motifs in each sample, and 4) fraction of aberrant fragments and average nucleotide frequencies surrounding fragment ends in each sample (GALYFRE). We trained and applied random forest machine learning models to analyze each set of these fragmentation features, as well as their combination in plasma DNA WGS data. Additional details about the features used, model generation, sample processing and statistical analyses are described in Supplementary Appendix.

## Results

To evaluate biomarker performance for detection of malignant PCNs, we analyzed plasma samples from 99 eligible patients with PCNs referred for EUS (**Figure 1**). Median age of patients was 72 years (IQR, 62 to 77 years) and 59 patients (59.6%) were women (additional clinical characteristics in **Table 1**). The dominant lesion observed on EUS was pancreas cyst in 84 patients and pancreatic duct dilatation in 15 patients. 24 patients (24.2%) had multiple cysts and/or pancreatic duct dilatation observed at EUS. For patients with pancreatic duct dilatation, the median lesion size measured at EUS was 0.7 cm (IQR, 0.5 to 1.0 cm). For patients with pancreas cysts, the median lesion size measured at EUS was 2.5 cm (IQR, 2.0 to 3.7 cm). One or more additional features of concern such as septations, mural nodularity, solid component or thickened cyst wall were observed at EUS in 37 patients with cysts (44.0%; **Supplementary Table S1**). Cyst fluid was analyzed by cytology for 72 patients (72.7%), and 25 patients (34.7%) were reported as either positive for malignancy, suspicious for mucinous neoplasm or showing atypical cells. Cyst fluid was sent for PancreaSeq genomic analysis for 58 patients (58.6%) and 6 patients (10.3%) were reported as intermediate or high-risk. A fine needle biopsy of the solid component was obtained in 10 patients (10.1%), and biopsy showed high-grade dysplasia, malignancy or a suspicion for malignancy in 5 patients (50.0%). Median time of clinical follow-up from blood collection was 24.6 months (IQR, 17.9 to 29.7 months). During this period, patients received regular clinical care, with a median number of 9 office, telemedicine or procedure visits across clinical specialties (IQR, 5 to 13 visits) and a median number of 2 imaging studies (IQR 1 to 3 imaging studies) (**Supplementary Figure S1**).

**Table 1:** Clinical characteristics of 99 patients with pancreatic cystic neoplasms. Malignant PCN is defined as diagnosis of high-grade dysplasia or invasive carcinoma during clinical work-up and surveillance. Additional lesion characteristics and individual level data are provided in Supplementary Table S1.

| Variable | Malignant PCN<br>(*n* = 11) | Benign PCN<br>(*n* = 88) | Total<br>(*n* = 99) | p-value |
|---|---|---|---|---|
| Sex, *n* (%) |  |  |  | 0.19 |
| Male | 2 (18.2) | 38 (43.2) | 40 (40.4) | - |
| Female | 9 (81.8) | 50 (56.8) | 59 (59.6) | - |
| Median age, years (IQR) | 72 (65, 77) | 70 (62, 77) | 72 (62, 77) | 0.84 |
| Age by Decade, *n* (%) |  |  |  | - |
| Under 50 years | 2 (18.2) | 8 (9.1) | 10 (10.1) | - |
| 50 to 59 years | 0 (0.0) | 9 (10.2) | 9 (9.1) | - |
| 60 to 69 years | 2 (18.2) | 24 (27.3) | 26 (26.3) | - |
| 70 to 79 years | 5 (45.5) | 34 (38.6) | 39 (39.4) | - |
| 80 years and over | 2 (18.2) | 13 (14.8) | 15 (15.2) | - |
| Race, *n* (%) |  |  |  | 1.0 |
| White | 11 (100.0) | 84 (95.5) | 95 (96.0) | - |
| Black | 0 (0.0) | 2 (2.3) | 2 (2.0) | - |
| Asian | 0 (0.0) | 2 (2.3) | 2 (2.0) | - |
| Ethnicity, *n* (%) |  |  |  | 1.0 |
| Hispanic or Latino | 0 (0.0) | 2 (2.3) | 2 (2.0) | - |
| Not Hispanic or Latino | 11 (100.0) | 86 (97.7) | 97 (98.0) | - |
| Lesion Type on EUS, *n* (%) |  |  |  | - |
| Pancreatic duct dilatation | 1 (9.1) | 14 (15.9) | 15 (15.2) | 1.0 |
| Cyst < 3 cm | 3 (27.3) | 47 (53.4) | 50 (50.5) | 0.08 |
| Cyst ≥ 3 cm | 7 (63.6) | 27 (30.7) | 34 (34.3) | - |
| High-risk cyst features on EUS* |  |  |  | 0.098 |
| Thickened cyst wall | 2 (20.0) | 2 (2.7) | 4 (4.8) | 0.068 |
| Septations | 1 (10.0) | 25 (33.8) | 26 (31.0) | 0.163 |
| Mural nodularity or solid component | 6 (60.0) | 4 (5.4) | 10 (11.9) | 0.0001 |
| Resection Type, *n* (%) |  |  |  | 0.69 |
| Pancreaticoduodenectomy | 2 (18.2) | 3 (3.4) | 5 (5.1) | - |
| Central Pancreatectomy | 0 (0.0) | 1 (1.1) | 1 (1.0) | - |
| Distal pancreatectomy | 4 (36.4) | 10 (11.4) | 14 (14.1) | - |
| Total pancreatectomy | 1 (9.1) | 0 (0.0) | 1 (1.0) | - |
| No surgery | 4 (36.4) | 74 (84.0) | 78 (78.8) | 0.0014 |
| Median plasma DNA concentration, ng/mL (IQR) | 5.8 (5.1, 14.3) | 7.1 (4.8, 10.5) | 6.9 (5.0, 10.9) | 0.96 |
| Vital Status |  |  |  | 0.0003 |
| Alive | 5 (45.5) | 82 (93.2) | 87 (87.9) | - |
| Dead due to pancreas cancer | 6 (54.5) | 0 (0.0) | 6 (6.1) | - |
| Dead due to other cause | 0 (0.0) | 5 (5.7) | 5 (5.1) | - |
| Dead due to unknown cause | 0 (0.0) | 1 (1.1) | 1 (1.0) | - |

Abbreviations: IQR, interquartile range; EUS, endoscopic ultrasound; SD, standard deviation.
Percentages may not add up to 100 due to rounding or multiple features observed in a single patient.
\*Some patients presented with multiple lesions. In this case, characteristics of the dominant lesion are described here. Statistical significance and proportions for high risk features observed at EUS are calculated for patients with pancreatic cysts (n=84) and excludes patients with PDD.

Eleven of 99 patients (11.1%) were found to have malignant PCNs during clinical workup and surveillance, including 2 patients with high-grade dysplasia, 1 patient with stage II solid pseudopapillary tumor, 3 patients with Stage I invasive carcinoma and 5 patients with Stage II invasive carcinoma. The diagnosis of malignant PCNs was confirmed upon surgical resection following EUS for 7 patients. The remaining 4 patients were diagnosed with malignancy by fine needle biopsy at EUS (3 patients during initial EUS and 1 patient at a follow-up EUS), and planned for neoadjuvant therapy prior to resection. Overall, based on recommendations of the multidisciplinary clinical team, 21 of 99 patients in this study underwent surgical resection and upon final histopathologic evaluation, 7 patients were confirmed to have malignant PCNs while 14 patients were diagnosed with benign lesions. During follow-up, 12 of 99 patients (12.1%) had died including 6 patients due to pancreatic cancer, and the remaining due to other causes.

For genome-wide plasma DNA fragmentation analysis, we first trained machine learning models to differentiate between patients with PDAC and healthy individuals, and then applied these models independently to patients with PCNs. In training and cross-validation, we analyzed 4 sets of features including GALYFRE, sequence end motifs, short-to-long fragment ratios and sequencing coverage using random forest models to detect pancreatic cancer in pre-treatment blood samples from 121 patients with PDAC (stages I-IV), and an equal number of age- and sex-matched healthy individuals. Clinical and sample characteristics of the training cohort are summarized in **Supplementary Table S2**. For differentiation between patients with PDAC and healthy individuals, we observed areas under the receiver operating characteristic curve (AUROCs) ranging from 0.88 to 0.96 for each set of fragmentomic features (**Figure 2a, Supplementary Table S3**). To evaluate whether a combination of these features may be more informative than each independent set, we combined all features into a single random forest model and observed an AUROC of 0.97. AUROCs were higher for detection of stage III (0.99) and stage IV (0.98), and lower for detection of stage I (0.94) and stage II cancers (0.97, **Figure 2b**). The combined plasma DNA fragmentation model achieved 90% sensitivity at a corresponding specificity of 88% for distinguishing patients with PDAC from healthy individuals.

**Figure 2:**
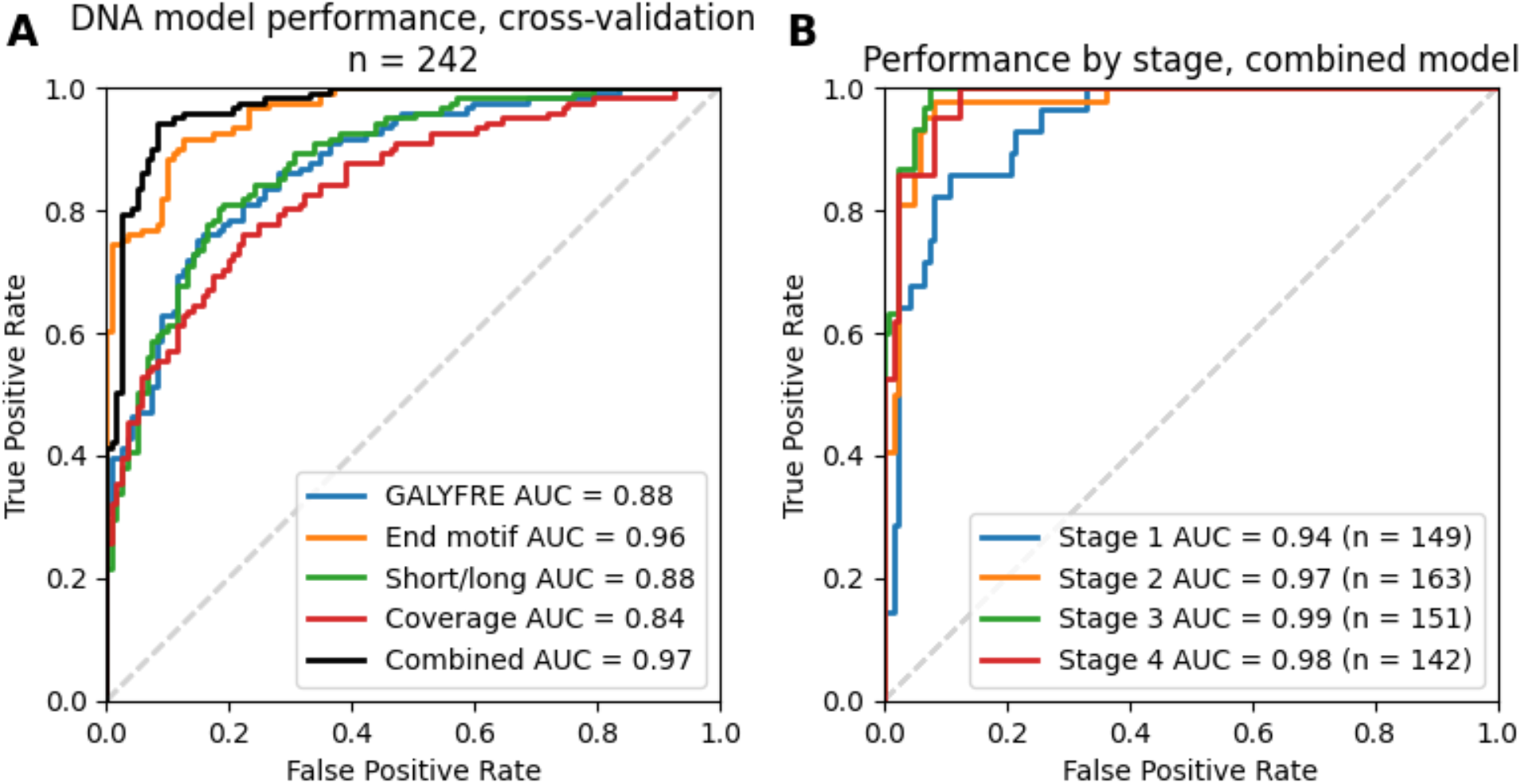
Evaluation of diagnostic performance for plasma DNA fragmentation analysis for detection of pancreatic cancer. **(A)** Receiver operating characteristic (ROC) curve demonstrating training and cross validation of different machine learning models to differentiate patients with PDAC from age- and sex-matched healthy controls using plasma DNA fragmentation analysis. Four sets of fragmentation features were evaluated including 4-mer sequence end motifs, variation in genome-wide sequencing coverage, variation in the ratio of short-to-long DNA fragments, and fraction of aberrant fragments as described in the GALYFRE model. A combined model using all four sets of fragmentation features was evaluated. **(B)** ROC curve demonstrating diagnostic performance of combined plasma DNA fragmentation analysis machine learning model separated by disease stage.

To evaluate if plasma DNA fragmentation analysis can help identify patients with malignant PCNs, we applied the locked plasma DNA fragmentation model to samples obtained from patients with PCNs referred for EUS. In patients with PCNs, we observed an AUROC of 0.67 using plasma DNA fragmentation analysis for detection of malignant PCNs (**Figure 3A, Supplementary Figure S2, Supplementary Table S4**). In the same patient cohort, using CA125, CA19-9, and CEA levels, we observed AUROCs of 0.54, 0.78 and 0.63 to differentiate patients with malignant vs. benign PCNs, respectively (**Figure 3B**). In a subgroup of patients with normal CA19-9 (below the clinical threshold), we observed an AUROC of 0.77 using plasma DNA fragmentation analysis (n=86 patients, **Supplementary Figure S3**). None of the three glycoproteins were correlated with each other, or with plasma DNA fragmentation scores, suggesting that each of these four biomarkers can be independently informative (**Supplementary Figure S4**).

**Figure 3:**
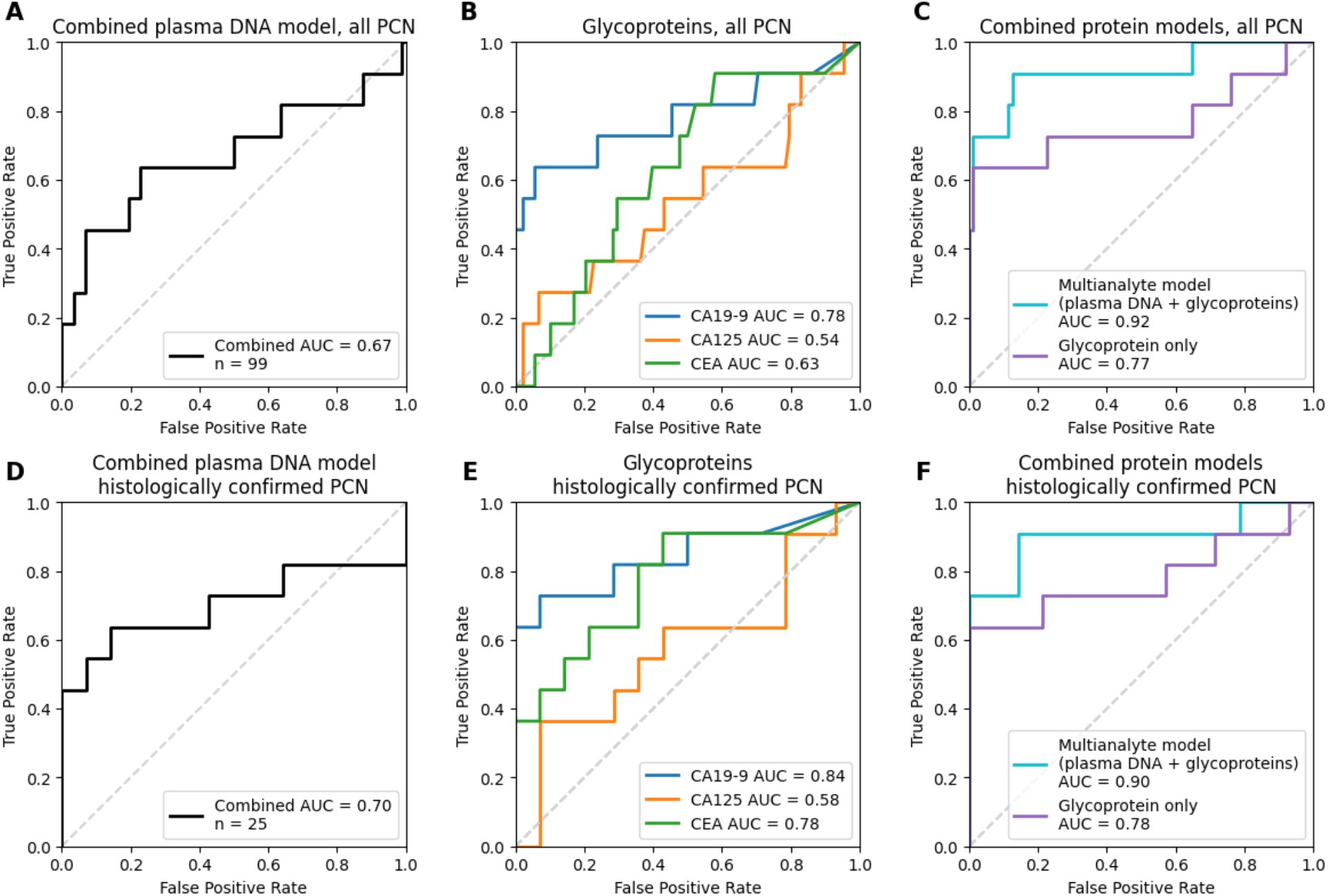
Evaluation of diagnostic performance for glycoprotein levels and plasma DNA fragmentation analysis in patients with pancreatic cystic neoplasms. **(A)** Performance of plasma DNA fragmentation model in all patients with PCNs (*n* = 99 patients) **(B)** Receiver operating characteristic (ROC) curves for three glycoprotein biomarkers analyzed individually: cancer antigen 125 (CA125), carbohydrate antigen 19-9 (CA19-9), carcinoembryonic antigen (CEA). Results show diagnostic performance in all patients with PCNs (*n* = 99 patients). **(C)** ROC curves generated using logistic regression to combine CA125, CA19-9, and CEA, with or without plasma DNA fragmentation, demonstrating diagnostic performance of a multianalyte blood test to differentiate patients with malignant and benign lesions amongst all patients with PCNs (*n* = 99 patients). **(D)** Performance of plasma DNA fragmentation model. Results show diagnostic performance in a subset of patients with PCN who received a definitive diagnosis (*n* = 25 patients). **(E)** ROC curves for four plasma biomarkers analyzed individually: CA125, CA19-9, CEA, and plasma DNA fragmentation scores in a subset of patients with PCNs with a histologically confirmed diagnosis (*n* = 25 patients). (F) Performance of logistic regression models with or without plasma DNA fragmentation in a subset of patients with PCNs with a histologically confirmed diagnosis (*n* = 25 patients).

Based on results from analysis of individual biomarkers, we evaluated the performance of a multianalyte approach combining CA125, CA19-9, CEA, and scores from the plasma DNA fragmentomic model using logistic regression. This approach showed an AUROC of 0.92 (95% CI of 0.785 to 0.995), higher than AUROCs observed for individual biomarkers (p=0.0053, 0.035, 0.0035 and 0.033 for comparisons with CA125, CA19-9, CEA, and plasma DNA fragmentation, respectively, **Figure 3C**). Using the combination of glycoproteins and plasma DNA fragmentation, the model surpassed 90% sensitivity (detected 10/11 patients with malignant PCNs) at a specificity of 87.5% (correctly identified 77/88 patients with benign PCNs) with a 47.6% positive predictive value (PPV, correct for 10/21 patients predicted as malignant) and a 98.7% negative predictive value (NPV, correct 77/78 patients predicted as malignant) for the detection of malignant PCNs in this cohort (at a model score threshold of 0.0965, **Supplementary Figure S2**). In contrast, individual biomarkers achieved 90% sensitivity at thresholds corresponding to specificity of 12.5% to 42% and PPV of 11.5% to 16.4%. When all 4 blood biomarkers were analyzed together with patient age, sex, lesion size and location of the lesion, using logistic regression, we observed an AUROC of 0.94 (**Supplementary Figure S5**). Consistent with the ROC curve, median multianalyte logistic regression scores in patients with malignant PCNs were 0.60 (*n* = 11, IQR 0.18 to 1.00), higher than patients with benign PCNs (*n* = 88, median 0.022, IQR 0.0073 to 0.073, *p* = 7.4x10^-6^), and higher than patients who underwent surgical resections for benign PCNs (*n* = 14, median 0.026, IQR 0.014 to 0.071, *p* = 7.5x10^-4^; **Supplementary Figure S6**).

We performed additional subgroup analysis and applied the same threshold for combined biomarker model score as described above (0.0965). In the subset of patients with pancreatic cysts of any size and no additional high-risk features on EUS (n=47), we observed 66% sensitivity (detected 2/3 patients with malignant PCNs) at 86.4% specificity (correctly identified 38/44 patients with benign PCNs), 97.4% NPV (correct for 38/39 patients predicted as benign), and 25.0% PPV (correct for 2/8 patients predicted as malignant). For patients with cysts and high-risk features on EUS (n=37), we observed 100% sensitivity (7/7 patients with malignant PCNs) at 86.7% specificity (26/30 patients with benign PCNs), 100%% NPV (correct for 26/26 patients predicted as benign) and 63.6% PPV (correct for 7/11 patients predicted as malignant). For patients with pancreatic duct dilatation (n=15), we observed 100% sensitivity (1/1 patients with malignant PCN) at 92.9% specificity (13/14 patients with benign PCNs), 100% NPV (correct for 13/13 patients predicted as benign) and 50.0% PPV (correct for 1/2 patients predicted as malignant). In the subset of patients with histologically confirmed diagnoses (11 patients with malignant PCNs vs. 14 patients with resection-confirmed benign PCNs), we observed an AUROC of 0.70 for plasma DNA fragmentation analysis (**Figure 3D, Supplementary Table S5**). Using CA125, CA19-9, and CEA levels, we observed AUROCs of 0.58, 0.84 and 0.78 to differentiate patients with malignant vs. benign PCNs, respectively (**Figure 3E**). In the same cohort, we observed an AUROC of 0.90 using combined analysis of plasma DNA, CA19-9, CA125 and CEA for detection of malignant PCNs (**Figure 3F**). Using the same model threshold as earlier, we observed 90.9% sensitivity (10/11 patients with malignant PCNs) at 85.7% specificity (12/14 patients with benign PCNs), 92.3% NPV (correct for 12/13 patients predicted as benign) and 83.3% PPV (correct 10/12 patients predicted as malignant; **Supplementary Figure S7**).

## Discussion

For patients with PCNs undergoing clinical evaluation, there are no well-established blood tests for detection of high-grade dysplasia or cancer. In this study, we investigated whether blood tests incorporating circulating tumor DNA can aid detection of malignancy, including high grade dysplasia, in patients with PCNs recommended for endoscopic ultrasound. We found that results of a multianalyte approach that combines plasma DNA fragmentation analysis in blood with CA19-9, CA125 and CEA may complement current diagnostic tools available for detection of malignant PCNs. Individually, CA19-9, CA125, CEA, and plasma DNA fragmentation analysis showed limited accuracy (54% to 78%). However, a multianalyte blood test combining plasma DNA fragmentation analysis with circulating glycoproteins showed an accuracy of 92% for detection of malignant PCNs in patients referred for EUS.

There are multiple points in the current diagnostic pathway for PCNs where such a blood test could be informative, and intended use of the test can help determine the appropriate quantitative thresholds. One approach is to improve selection of patients undergoing EUS to optimize the use of clinical resources and to minimize invasive testing in patients unlikely to benefit, without missing malignant PCNs (**Figure 4**). For this application, an appropriate threshold can be chosen for a rule-out blood test to achieve high sensitivity, high negative predictive value and a positive predictive value that improves over current standard of care. In our study, combined testing of glycoprotein and plasma DNA biomarkers achieved 90% sensitivity (detected 10 of 11 patients with malignant lesions) at a PPV of 47.6% such that 10 of 21 patients above this threshold were eventually diagnosed with a malignant PCN. In comparison, using the current standard of care for selecting patients for EUS in our study, 11% of patients (11 of 99 patients) were diagnosed with a malignant PCN, comparable to similarly reported rates in literature. In patients with pancreatic cysts of any size without additional concerning features, the blood test showed a PPV of 25.0% (2 of 8 patients), while 6.4% (3 of 47 patients) were diagnosed with a malignant PCN during the standard course of management. Although additional validation and larger studies are needed to refine our estimates of PPV, these results suggest similar rates of detection of malignant PCN could be achieved with a blood test-first approach in this cohort with 5-fold fewer patients undergoing EUS for benign lesions.

**Figure 4:**
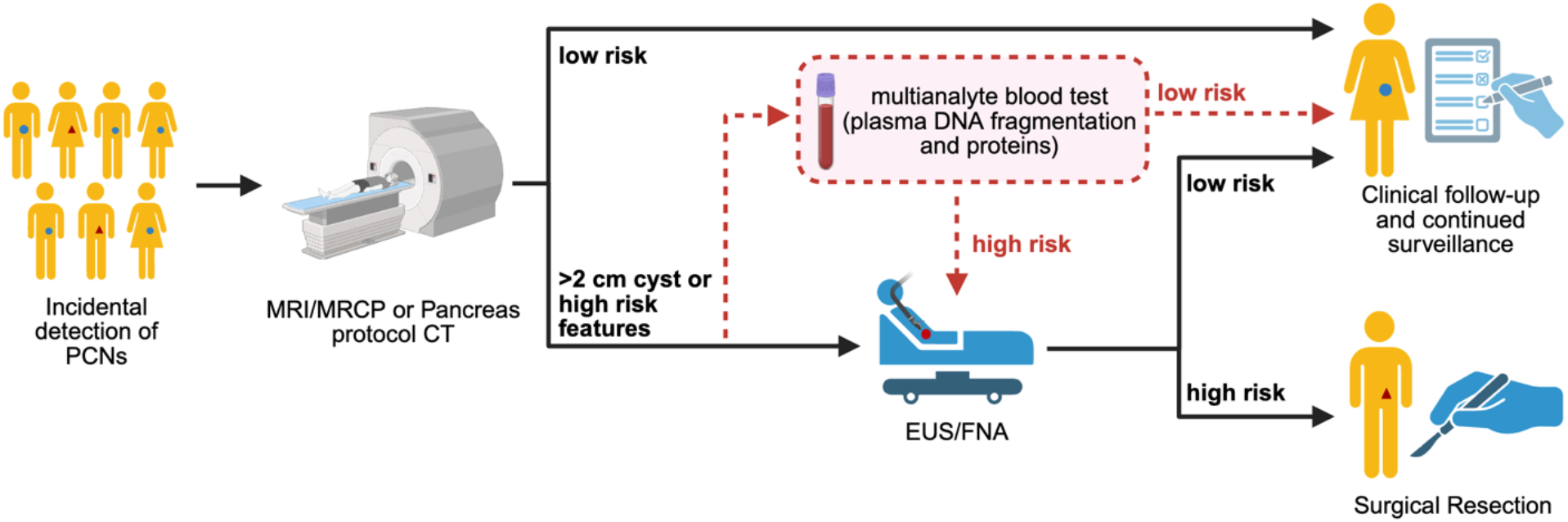
Proposed application of a multianalyte blood test combining plasma DNA fragmentation analysis and glycoprotein biomarkers. Based on observations in the current study and pending further validation, a multianalyte blood test could be applied to further stratify patients with incidental PCNs referred for EUS after imaging. Clinical practice patterns for selection of patients with PCNs for EUS differ between institutions. Our results show a blood-test first approach particularly for patients with PCNs where cyst size is the primary driver of EUS referral could improve overall positive predictive value. With this approach, patients who are determined to be low risk based on the blood test result can skip EUS and return to clinical follow-up. In contrast, patients who are determined to be high risk on the blood test can undergo EUS and continue downstream risk assessment and management as per standard of care.

Using non-invasive blood testing as a first-pass test to improve patient selection for invasive diagnostic procedures such as EUS and FNA is analogous to the use of non-invasive prenatal testing using maternal plasma cell-free DNA sequencing for screening of Down syndrome in pregnant women^36^. Compared to the prior standard of care for screening with serum biochemical assays and nuchal translucency measurement, cell-free DNA sequencing achieved similarly high levels of sensitivity but improved positive predictive value from 4.2% to 45.5%, reducing the number of women unnecessarily exposed to invasive amniocentesis or chorionic villus sampling and the risk of complications. Similarly, for patients with higher risk PCNs particularly those with large cysts but no other concerning findings, our results suggest that a blood test-first approach could help optimize resource use and selection of patients most likely to benefit from EUS.

There are several limitations of this study which need to be addressed in future work. One key limitation is that clinical practice patterns for management of PCNs and referral for EUS vary between institutions. Our study has a limited sample size of 99 patients with PCNs undergoing EUS and reflects the results of multidisciplinary evaluation at a single institution. In particular, we note ∼32% of our patients had pancreatic cysts smaller than 3 cm in size and no additional high-risk features observed at EUS, although importantly two of these patients were diagnosed with a malignant PCN. While we have found evidence that a multianalyte blood test with plasma DNA fragmentation and glycoprotein analysis could improve patient selection for EUS, a larger multicenter study is needed to identify the appropriate diagnostic thresholds, improve estimates of sensitivity and PPV, and perform more effective subgroup analyses. Another limitation of this study is that a negative EUS does not definitively rule out malignancy and it is possible that some patients we have classified as having benign PCNs may develop a malignancy in the future. However, these patients did not develop PDAC during a median clinical follow-up of 24.6 months, and the results of the multianalyte blood test remained robust when we limited analysis to a subset of patients with definitive tissue diagnoses. Finally, this study of plasma DNA and glycoprotein analysis was limited to patients with PCNs, and our findings cannot be extended without further evaluation to other patients at high risk of developing pancreatic cancer such as those with chronic pancreatitis, strong family history of pancreatic cancer, and new-onset diabetes.

In summary, we have found that a multianalyte blood test that incorporates plasma DNA fragmentation analysis with glycoprotein biomarkers can serve as a diagnostic aid for detection of high-grade dysplasia or carcinoma in patients with PCNs. Our results suggest this blood test can improve selection of patients at the highest risk of harboring a malignant PCN who are likely to benefit from further invasive testing such as EUS or surgical resection. While promising, larger multicenter studies of patients with pancreatic cystic neoplasms are needed to validate our findings, and to establish the best strategy to incorporate this blood-based approach together with other available modalities for cancer detection.

## Data Availability

Sequencing data included in this manuscript will be deposited to a controlled access public database once the manuscript is accepted at a peer-review journal. All other data produced in the present work are contained in the manuscript.

## ACKNOWLEDGEMENTS

The authors wish to thank the following individuals, groups, and resources for their support: the nurses and staff of the UW Health Pancreas Cancer Prevention Clinic and the University Hospital Ambulatory Procedure Center for their assistance with sample collection; Dr. David Yang, Mr. Kevin Grant, and the UW Health Clinical Laboratory for providing immunoassay services; the UW Clinical Research Data Service for their contributions to clinical data acquisition; and Mr. Will Suter, Ms. Judy Wang, Ms. Hellen Nginga, and UW Carbone Cancer Center BioBank, supported by grant P30 CA014520 from the NCI of the NIH, for sample collection and processing.

## Abbreviation list

AUROC: area under the receiver operating characteristic curve
CA125: cancer antigen 125
CA19-9: carbohydrate antigen 19-9
CEA: carcinoembryonic antigen
EUS: endoscopic ultrasound
HGD: high grade dysplasia
IQR: interquartile range
MRI: magnetic resonance imaging
PCN: Pancreatic Cystic Neoplasms
PCPC: Pancreatic Cancer Prevention Clinic
PDAC: pancreatic ductal adenocarcinoma
PPV: positive predictive value
ROC: receiver operating characteristic
SD: standard deviation
WGS: whole genome sequencing

