## Supplementary Appendix for "Plasma DNA fragmentomic analysis for evaluation of patients with pancreatic cystic neoplasms"

### Supplementary Methods

Plasma processing and DNA extraction

For patients with PCNs, blood samples were collected in $K_2$ ethylenediaminetetraacetic acid (EDTA) tubes, plasma was isolated within one hour of sample collection, and stored at –80 ºC until DNA extraction. DNA was extracted from 800 µL to 1 mL of plasma using the MagMax Cell-Free DNA Isolation Kit (ThermoFisher Scientific, Waltham, Massachusetts, USA). Plasma DNA samples were quantified using the 4200 TapeStation automated electrophoresis system (Agilent Technologies, Santa Clara, California, USA), and stored at –20 ºC until library preparation.

Whole genome sequencing

Sequencing libraries were prepared using the ThruPLEX DNA-Seq HV kit (Takara Bio, Kusatsu, Shiga, Japan), following manufacturer's instructions, and quantified using the 4200 TapeStation system (Agilent) and the Qubit Flex fluorometer (ThermoFisher). WGS was performed on NovaSeq X Plus system (Illumina Inc., San Diego, California, USA), generating 50 bp paired-end reads. After on-board FASTQ conversion and sample demultiplexing, sequencing data were aligned to human genome build hg19 using BWA-MEM.[1] SAMtools was used to mark duplicate reads and calculate alignment statistics.[2] Only reads that were non-duplicate, properly paired, and with a minimum mapping quality of 60 were used for further analysis. For the training and validation cohort, we obtained a median depth of sequencing coverage of 1.34× (IQR, 1.08× to 1.61×) for patients with cancer and 1.61× (IQR, 1.34× to 2.09×) for healthy individuals. For patients with PCNs, we achieved median depth of sequencing coverage of 1.38× (IQR, 1.08× to 1.59×).

Computational analysis of plasma DNA fragmentation patterns

We trained and validated random forest machine learning models based on plasma DNA fragmentation features to specifically detect pancreatic cancer. Four sets of fragmentomic features were evaluated:

GALYFRE, genome-wide variation in short-to-long fragment ratios, nucleotide motifs at plasma DNA fragment ends, and genome-wide variation in normalized sequencing coverage.

For GALYFRE, feature calculations and feature selection were previously established and detailed descriptions are included in the original publication.[3] Genome-wide variation in short-to-long fragment ratios features were calculated using code provided in the original publication.[4] Nucleotide motifs at plasma DNA fragment ends were calculated using BEDTools v2.31.1 [5]. Genome-wide variation in GC-normalized sequencing coverage was calculated for 1 megabase bins using ichorCNA.[6]
For each set of features, a random forest machine learning model was trained and cross-validated in healthy individuals and PDAC patients to train a model based specifically on pancreatic cancer. In addition to training a model on each feature set individually, we trained a model by combining all feature sets, allowing the choice of the most informative combination of features. These machine learning model assigns an overall score to each sample that represents its similarity in fragmentation patterns to samples from patients with cancer versus those from healthy individuals, ranging from 0 (least likely to be from a patient with cancer) to 1 (most likely to be from a patient with cancer). All models were generated using Scikit-learn.[7] The combined model was locked and we independently assessed the diagnostic performance of this model in in patients with PCNs.

<u>Analysis of plasma protein biomarkers</u>

All three proteins (CA19-9, CA125 and CEA) were measured using the chemiluminescent immunoassay performed on the ARCHITECT *i*2000SR analyzer (Abbott Laboratories, Green Oaks, Illinois, USA).

<u>Logistic regression model</u>

To incorporate plasma DNA analysis with protein biomarkers, logistic regression was performed on samples from patients with PCNs. Protein values were normalized for interpretability based on clinical thresholds. For the multianalyte regression, four features were incorporated: combined plasma DNA model score, CA125, CA19-9, and CEA. The logistic regression model was fit using statsmodels[8], with an added constant and no penalty. A model assessing performance of the three glycoproteins (CA19-9, CA125, and CEA) was produced in the same manner. To assess the effect of including clinical data, an additional logistic regression model was constructed from the plasma features (DNA model score, CA125, CA19-9, and CEA) plus the age and sex of the patient and the lesion type and size in centimeters. Categorical variables were one-hot encoded. The model was fit with an added constant and no penalty.

Statistical methods

Descriptive statistics are described as frequency for categorical variables and median with interquartile range (IQR) for continuous variables. The Chi-square test was used to compare categorical variables, and the Mann-Whitney *U* test was used for continuous variables. Median follow-up with IQR was calculated using the reverse Kaplan-Meier estimator. To evaluate performance of individual protein biomarkers, plasma DNA fragmentation analysis, as well as the multianalyte logistic regression, we assessed the area under the receiving operator characteristic curve (AUROC). To assess the statistical significance of AUROC values, the Mann-Whitney *U* test was used.[9] To compare the AUROC performance between single biomarkers and combined models, the DeLong test was used[10]. An alternative analysis of classification accuracy was performed by assigning predicted classes using each individual biomarker or their combined logistic regression score based on the value corresponding to 90% sensitivity. All *p*-values are two-sided, with a value $\leq 0.05$ considered statistically significant. All analyses were performed using SciPy[11] with Python, version 3.13. Study data were collected and managed using Research Electronic Data Capture (REDCap) tools hosted by the Institute for Clinical and Translational Research at the University of Wisconsin–Madison.[12]

### Supplementary Figures

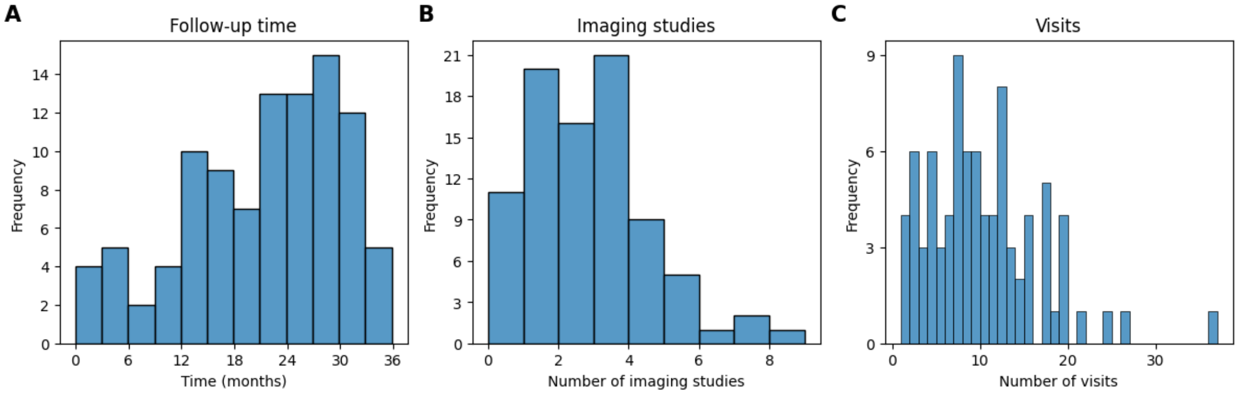

**Supplementary Figure S1:** Follow-up of patients with PCNs. **A.** Histogram of follow-up time for each patient in the study. **B.** Histogram of number of imaging studies for each patient during follow-up. Patients with confirmed malignant pathology were excluded. **C.** Histogram of clinical visits tracked for each patient during follow-up. Patients with confirmed malignant pathology were excluded.

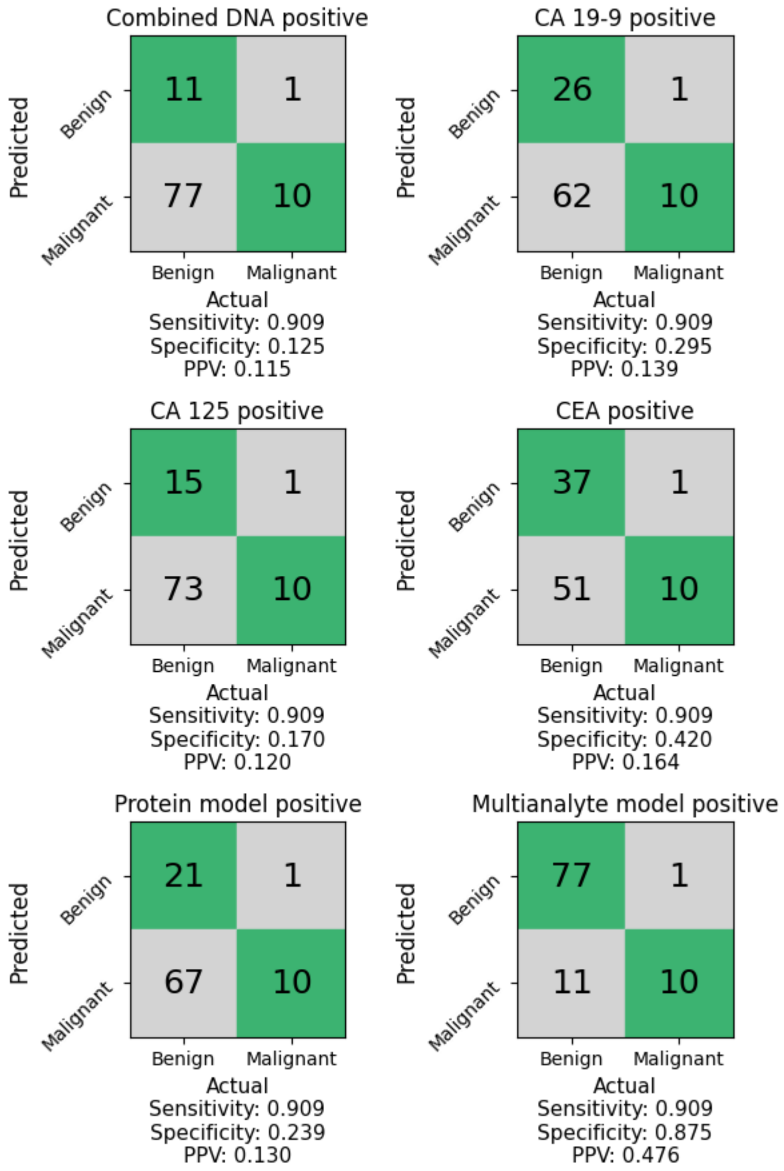

**Supplementary Figure S2:** Confusion matrices for individual evaluation of plasma DNA fragmentation, glycoprotein biomarkers, and combined multianalyte regression at 90% sensitivity. Predicted and actual classification of malignant pathology versus benign pathology are shown. A multianalyte model incorporating plasma DNA fragmentation as well as glycoprotein biomarkers demonstrates the highest specificity and PPV.

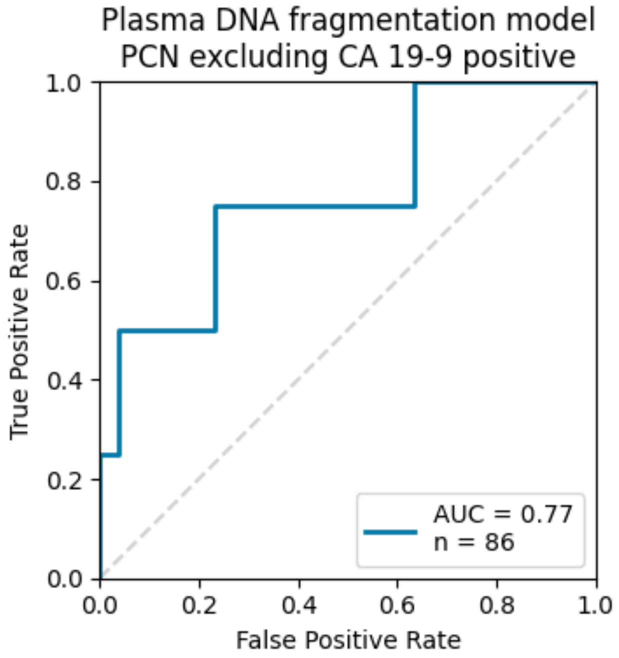

**Supplementary Figure S3:** Evaluation of diagnostic performance of the plasma DNA fragmentation model in A subset of patients with normal CA19-9 levels (below the clinical threshold of 37 U/mL).

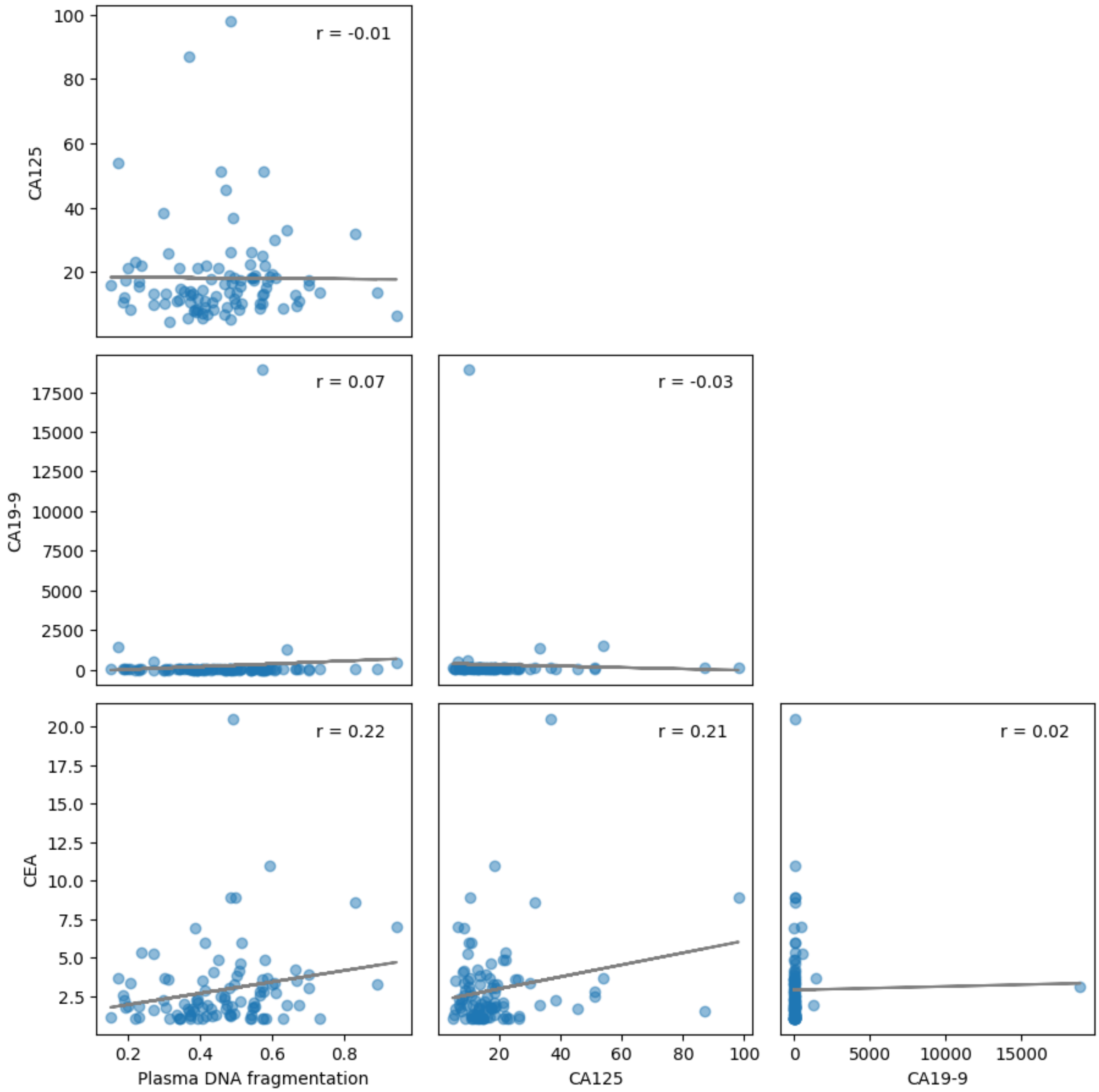

**Supplementary Figure S4:** Evaluation of correlation between plasma DNA fragmentation score and glycoprotein biomarkers. The linear least-squares regression line is in grey, and the Pearson correlation coefficient is indicated for each pair of biomarkers.

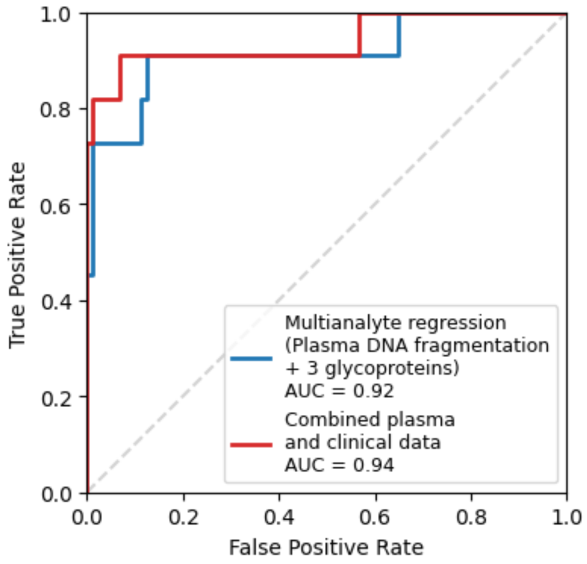

**Supplementary Figure S5:** Evaluation of differences in diagnostic performance using the multianalyte blood test when additional clinical variables were included in logistic regression (age, sex, lesion size and location of the lesion).

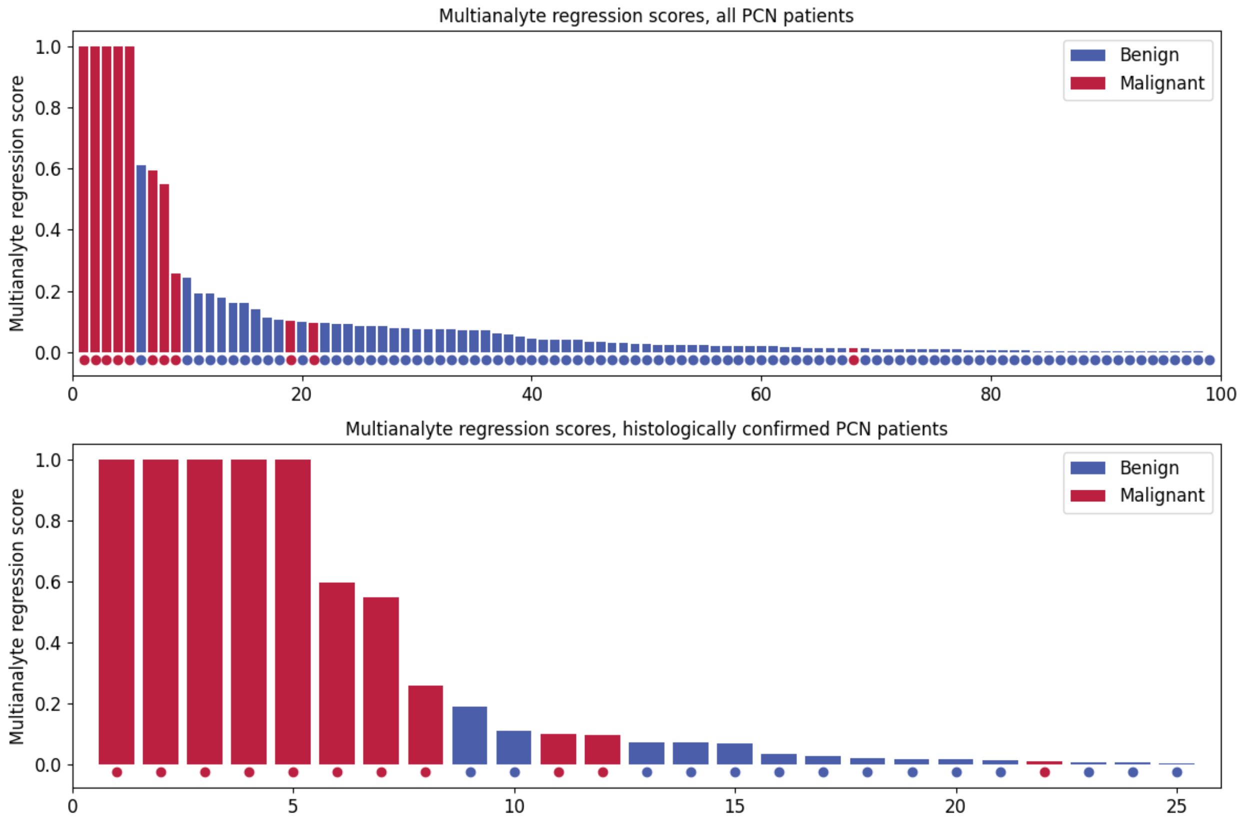

**Supplementary Figure S6:** Multianalyte regression model scores for patients with incidental pancreas lesions. Top: all patients with pancreatic cysts (*n* = 99). Bottom: histologically confirmed subset of patients who initially presented with pancreatic cysts (*n* = 25). This subset includes 11 patients diagnosed with malignant pathology (upon fine needle aspiration, biopsy or surgical resection) and 14 patients with benign pathology (confirmed histologically after surgical resection).

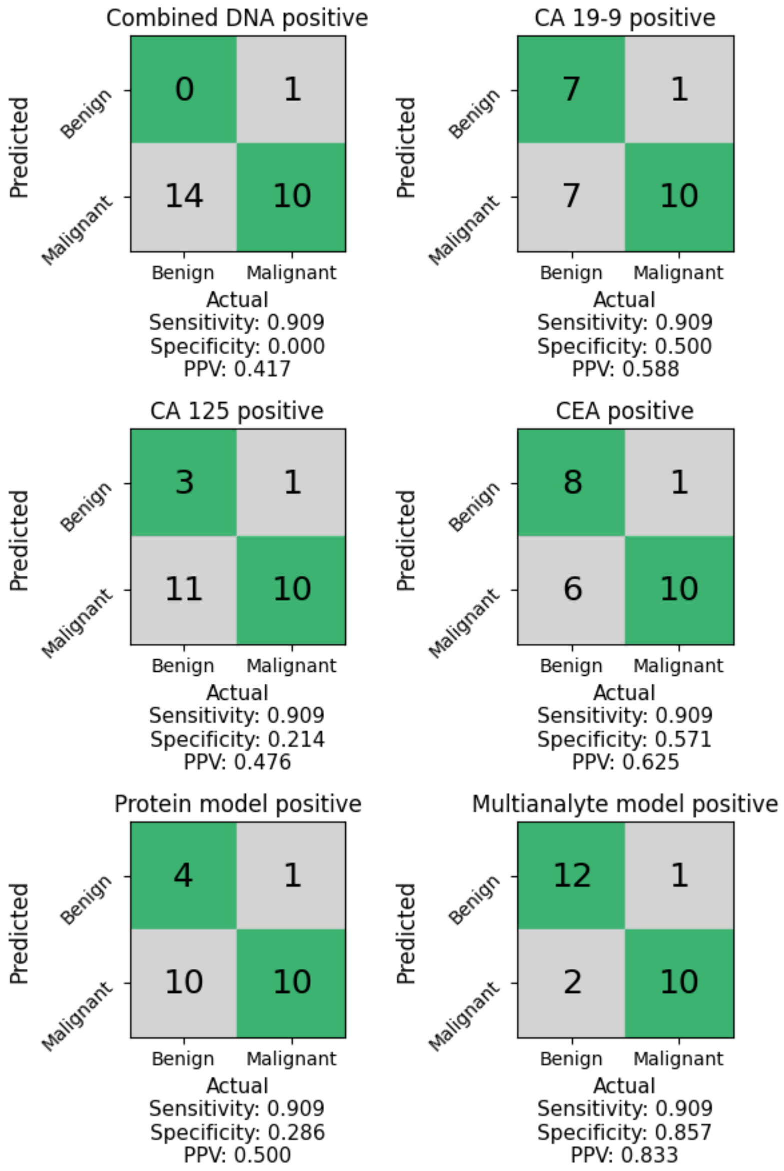

**Supplementary Figure S7**: Confusion matrices for individual evaluation of glycoprotein biomarkers, plasma DNA fragmentation and combined multianalyte regression, in patients with histologically confirmed diagnoses (n=25). Predicted and actual classification of malignant pathology versus benign pathology are shown. A combination of the plasma DNA model, CA19-9, and CA125 demonstrates the highest specificity at a threshold for 90% sensitivity.

### Supplementary Tables

**Supplementary Table S1:** Additional clinical and pathologic information for the patients with incidental pancreas lesions.

| Patient ID | Sex | Pathology | Lesion Type* | Lesion Size at EUS (in cm) | Lesion Location | Additional EUS Lesion Features | Surgery | Vital Status |
|---|---|---|---|---|---|---|---|---|
| P2 | Female | Benign | Cyst | 3.0 | Uncinate | | Yes | Alive |
| P3 | Male | Benign | Cyst | 3.4 | Head | S | No | Alive |
| P5 | Female | Benign | PDD | 0.8 | Body | | No | Alive |
| P6 | Female | Benign | Cyst | 2.3 | Tail | S | No | Alive |
| P7 | Female | Benign | Cyst | 0.8 | Tail | | No | Alive |
| P8 | Female | Benign | Cyst | 4.2 | Uncinate | S | No | Alive |
| P9 | Male | Benign | Cyst | 2.3 | Body | | No | Alive |
| P11 | Male | Benign | Cyst | 5.0 | Body | | No | Alive |
| P13 | Female | Benign | Cyst | 10.0 | Body | S, W | Yes | Alive |
| P14 | Female | Malignant | Cyst | 5.8 | Head | N | Yes | Dead (pancreas cancer) |
| P16 | Female | Benign | Cyst | 2.3 | Neck | S | No | Alive |
| P18 | Female | Benign | Cyst | 4.0 | Body | | No | Alive |
| P19 | Female | Malignant | Cyst | 4.5 | Tail | S, SC | Yes | Alive |
| P22 | Female | Benign | Cyst | 2.9 | Body | | No | Alive |
| P23 | Female | Benign | Cyst | 2.1 | Tail | | No | Alive |
| P24 | Male | Benign | Cyst | 1.7 | Body | S | No | Alive |
| P25 | Female | Benign | PDD | 0.5 | Body | | Yes | Alive |
| P29 | Male | Benign | Cyst | 3.6 | Tail | N | No | Alive |
| P31 | Male | Benign | Cyst | 4.8 | Body | | No | Alive |
| P33 | Female | Benign | Cyst | 1.2 [a] | Body | | No | Alive |
| P34 | Female | Benign | Cyst | 4.0 | Head | S | No | Alive |
| P35 | Female | Benign | Cyst | 1.6 | Uncinate | S | No | Alive |
| P36 | Female | Benign | Cyst | 1.9 | Head | | No | Alive |
| P37 | Female | Benign | Cyst | 2.5 | Body | | No | Alive |
| P38 | Male | Benign | Cyst | 6.7 | Body | | No | Alive |
| P39 | Female | Benign | Cyst | 2.4 | Body | | No | Alive |
| P41 | Male | Benign | PDD | 1.3 | Tail | | No | Dead (other cause) |
| P43 | Male | Benign | Cyst | 1.8 [a] | Tail | | No | Alive |
| P44 | Female | Benign | Cyst | 2.4 | Head | | No | Alive |
| P45 | Female | Benign | Cyst | 0.9 [a] | Tail | | No | Alive |
| P47 | Male | Benign | Cyst | 2.4 | Uncinate | | No | Dead (other cause) |
| P49 | Female | Malignant | Cyst | 3.1 | Body | | Yes | Alive |
| P50 | Male | Benign | Cyst | 2.8 | Neck | S | No | Dead (other cause) |
| P51 | Female | Benign | Cyst | 2.0 | Head | | No | Alive |

| | | | | | | | | |
|---|---|---|---|---|---|---|---|---|
| P53 | Male | Benign | Cyst | 3.6 | Tail | | No | Alive |
| P54 | Male | Benign | PDD | 0.6 | Body | | No | Alive |
| P55 | Female | Benign | Cyst | 5.0 | Neck | SC | No | Alive |
| P56 | Female | Benign | Cyst | 5.0 | Head | S | No | Alive |
| P58 | Female | Benign | PDD | 1.6 | Body | | No | Alive |
| P59 | Female | Malignant | Cyst | 3.8 | Tail | W, SC | Yes | Alive |
| P60 | Female | Benign | Cyst | 1.2 | Body | | No | Alive |
| P61 | Male | Benign | Cyst | 3.8 | Head | | No | Alive |
| P62 | Female | Benign | Cyst | 2.7 | Tail | S | No | Alive |
| P63 | Male | Benign | Cyst | 1.5 [a] | Tail | | No | Alive |
| P64 | Female | Benign | PDD | 1.6 | Body | | No | Dead (other cause) |
| P65 | Male | Benign | Cyst | 3.5 | Head | | No | Dead (other cause) |
| P66 | Male | Benign | Cyst | 2.0 | Body | | No | Alive |
| P67 | Male | Benign | PDD | 0.7 | Neck | | Yes | Alive |
| P68 | Male | Benign | Cyst | 2.2 | Body | W | Yes | Alive |
| P70 | Male | Malignant | Cyst | 4.8 | Uncinate | W | Yes | Alive |
| P71 | Female | Benign | Cyst | 2.1 | Body | | No | Alive |
| P73 | Male | Benign | Cyst | 4.0 | Tail | | No | Alive |
| P75 | Male | Benign | Cyst | 2.8 | Head | S | No | Alive |
| P76 | Female | Malignant | Cyst | 3.0 | Head | SC | No | Dead (pancreas cancer) |
| P77 | Female | Benign | Cyst | 2.0 | Head | | No | Alive |
| P78 | Female | Benign | Cyst | 2.5 | Tail | S | No | Alive |
| P80 | Female | Benign | Cyst | 2.0 | Tail | | Yes | Alive |
| P81 | Female | Benign | Cyst | 3.0 | Uncinate | | No | Alive |
| P82 | Male | Benign | Cyst | 2.1 | Tail | | No | Alive |
| P83 | Female | Malignant | Cyst | 2.3 | Body | | Yes | Alive |
| P85 | Female | Malignant | PDD | 0.5 | Neck | | No | Dead (pancreas cancer) |
| P86 | Male | Benign | Cyst | 1.6 | Tail | S | Yes | Alive |
| P87 | Male | Benign | Cyst | 1.8 | Neck | | No | Alive |
| P88 | Female | Benign | Cyst | 3.0 | Uncinate | | No | Alive |
| P89 | Male | Benign | Cyst | 3.0 | Tail | S | No | Alive |
| P90 | Female | Malignant | Cyst | 2.5 | Body | SC | No | Dead (pancreas cancer) |
| P91 | Female | Benign | Cyst | 7.0 | Body | S | No | Alive |
| P92 | Male | Benign | Cyst | 3.6 | Body | S | No | Alive |
| P93 | Female | Benign | Cyst | 4.5 | Head | | Yes | Alive |
| P94 | Female | Benign | PDD | 0.4 | Head | | No | Alive |
| P96 | Female | Benign | Cyst | 3.5 | Body | S | No | Alive |
| P97 | Female | Benign | PDD | 0.8 | Body | | No | Alive |
| P99 | Male | Benign | Cyst | 2.4 | Tail | | No | Alive |
| P100 | Female | Benign | Cyst | 2.0 | Head | | No | Alive |
| P102 | Female | Benign | Cyst | 2.2 | Head | S | No | Alive |
| P103 | Male | Benign | Cyst | 2.0 | Body | | No | Alive |

| | | | | | | | | |
|---|---|---|---|---|---|---|---|---|
| P104 | Female | Malignant | Cyst | 3.0 | Body | SC | No | Dead (pancreas cancer) |
| P107 | Male | Benign | PDD | 0.4 | Neck | | Yes | Alive |
| P119 | Female | Benign | PDD | 1.1 | Neck | | Yes | Alive |
| P120 | Female | Benign | Cyst | 4.0 | Head | | No | Alive |
| P241 | Female | Benign | Cyst | 1.8 | Neck | S | No | Alive |
| P242 | Female | Benign | Cyst | 6.4 | Uncinate | | No | Alive |
| P245 | Female | Benign | PDD | 0.5 | Body | | Yes | Alive |
| P247 | Female | Benign | Cyst | 2.1 | Body | | No | Alive |
| P249 | Female | Benign | Cyst | 2.0 | Head | | No | Alive |
| P251 | Female | Benign | Cyst | 2.3 | Body | SC | Yes | Alive |
| P253 | Female | Benign | Cyst | 1.9 | Tail | | Yes | Alive |
| P255 | Male | Benign | Cyst | 2.3 | Head | S | Yes | Alive |
| P256 | Male | Malignant | Cyst | 2.0 | Head | | Yes | Dead (pancreas cancer) |
| P257 | Male | Benign | Cyst | 4.0 | Uncinate | S | No | Alive |
| P258 | Female | Benign | Cyst | 2.0 | Head | | No | Alive |
| P259 | Male | Benign | Cyst | 4.0 | Body | S | No | Alive |
| P261 | Male | Benign | Cyst | 2.6 | Body | S | No | Alive |
| P262 | Male | Benign | PDD | 0.6 | Body | | No | Alive |
| P263 | Female | Benign | Cyst | 2.5 | Tail | S | No | Alive |
| P265 | Male | Benign | Cyst | 2.8 | Uncinate | SC | No | Alive |
| P266 | Male | Benign | PDD | 0.9 | Head | | No | Dead (unknown cause) |
| P267 | Male | Benign | Cyst | 1.8 | Head | | No | Alive |
| P268 | Male | Benign | Cyst | 1.6 | Body | | No | Alive |

* Some patients presented with multiple lesions. In this case, the dominant lesion is noted.
[a] Lesion demonstrated a thickened or enhancing component on cross-sectional imaging.
Abbreviations: PDD, pancreatic duct dilatation; S, septations; N, mural nodularity; W, thickened cyst wall; SC, solid component

**Supplementary Table S2:** Summary of clinical characteristics of patients with PDAC and healthy individuals used to train a machine learning model for plasma DNA fragmentation analysis.

| Variable | PDAC ($n$ = 121) | Healthy ($n$ = 121) |
|---|---|---|
| Sex, *n* (%) | | |
| Male | 56 (46.3) | 56 (46.3) |
| Female | 65 (53.7) | 65 (53.7) |
| Median age, years (IQR) | 69.0 (61.0, 75.0) | 69.0 (61.0, 75.0) |
| Age by Decade, *n* (%) | | |
| Under 50 | 0 (0.0) | 0 (0.0) |
| 50 to 59 | 21 (17.4) | 21 (17.4) |
| 60 to 69 | 38 (31.4) | 38 (31.4) |
| 70 to 79 | 46 (38.0) | 48 (39.7) |
| 80 and over | 14 (11.6) | 12 (9.9) |
| Collection location, *n* (%) | | |
| UW–Madison Hospital | 55 (45.5) | 0 (0.0) |
| Royal Surrey Hospital | 66 (54.5) | 0 (0.0) |
| Community collection | 0 (0.0) | 121 (100.0) |
| AJCC Stage, *n* (%) | | |
| I | 28 (23.1) | |
| II | 42 (34.7) | |
| III | 30 (24.8) | |
| IV | 21 (17.4) | |
| Median plasma cfDNA concentration, ng/mL (IQR) | 9.2 (5.4, 22.7) | 6.5 (4.3, 9.9) |

Abbreviations: PDAC, pancreatic ductal adenocarcinoma; IQR, interquartile range; AJCC, American Joint Commission on Cancer; cfDNA, cell-free DNA.

**Supplementary Table S3:** Plasma DNA fragmentation scores for individual samples from patients with PDAC and healthy individuals used to train a machine learning model for plasma DNA fragmentation analysis.

| Patient ID | Sex | Status | AJCC Stage | GALYFRE score | End motif score | Short/ long score | Coverage score | Combined DNA score |
|---|---|---|---|---|---|---|---|---|
| M1109 | Male | PDAC | II | 0.750 | 0.958 | 0.824 | 0.521 | 0.767 |
| M1110 | Male | PDAC | IV | 0.675 | 0.922 | 0.734 | 0.632 | 0.794 |
| M1111 | Female | PDAC | II | 0.363 | 0.819 | 0.411 | 0.371 | 0.572 |
| M1112 | Male | PDAC | III | 0.606 | 0.922 | 0.621 | 0.490 | 0.666 |
| M1114 | Female | PDAC | IV | 0.951 | 0.901 | 0.740 | 0.669 | 0.791 |
| M1115 | Female | PDAC | II | 0.638 | 0.958 | 0.475 | 0.218 | 0.616 |
| M1116 | Male | PDAC | III | 0.916 | 0.870 | 0.943 | 0.906 | 0.928 |
| M1117 | Female | PDAC | II | 0.535 | 0.815 | 0.598 | 0.394 | 0.716 |
| M1121 | Female | PDAC | II | 0.507 | 0.818 | 0.694 | 0.325 | 0.646 |
| M1123 | Male | PDAC | I | 0.138 | 0.739 | 0.660 | 0.338 | 0.545 |
| M1127 | Female | PDAC | I | 0.307 | 0.704 | 0.283 | 0.282 | 0.402 |
| M1128 | Male | PDAC | I | 0.787 | 0.854 | 0.450 | 0.421 | 0.630 |
| M1129 | Female | PDAC | III | 0.920 | 0.895 | 0.789 | 0.780 | 0.846 |
| M1130 | Female | PDAC | II | 0.509 | 0.711 | 0.820 | 0.434 | 0.703 |
| M1133 | Female | PDAC | III | 0.921 | 0.884 | 0.863 | 0.633 | 0.803 |
| M1135 | Male | PDAC | II | 0.587 | 0.685 | 0.407 | 0.494 | 0.563 |
| M1136 | Male | PDAC | I | 0.521 | 0.732 | 0.639 | 0.685 | 0.683 |
| M1137 | Female | PDAC | IV | 0.486 | 0.841 | 0.510 | 0.226 | 0.598 |
| M1138 | Male | PDAC | II | 0.603 | 0.794 | 0.325 | 0.464 | 0.625 |
| M1140 | Male | PDAC | II | 0.640 | 0.846 | 0.389 | 0.372 | 0.636 |
| M1141 | Male | PDAC | III | 0.645 | 0.755 | 0.622 | 0.573 | 0.664 |
| M1143 | Female | PDAC | I | 0.368 | 0.840 | 0.367 | 0.475 | 0.630 |
| M1145 | Male | PDAC | IV | 0.792 | 0.797 | 0.790 | 0.494 | 0.714 |
| M1146 | Female | PDAC | II | 0.806 | 0.951 | 0.385 | 0.430 | 0.663 |
| M1147 | Female | PDAC | II | 0.920 | 0.834 | 0.939 | 0.847 | 0.880 |
| M1149 | Female | PDAC | IV | 0.790 | 0.797 | 0.626 | 0.564 | 0.746 |
| M1150 | Male | PDAC | II | 0.850 | 0.748 | 0.720 | 0.460 | 0.614 |
| M1152 | Male | PDAC | I | 0.542 | 0.933 | 0.426 | 0.307 | 0.630 |
| M1153 | Female | PDAC | IV | 0.526 | 0.890 | 0.579 | 0.644 | 0.736 |
| M1154 | Female | PDAC | III | 0.576 | 0.788 | 0.270 | 0.339 | 0.553 |
| M1159 | Male | PDAC | II | 0.704 | 0.968 | 0.428 | 0.279 | 0.597 |
| M1164 | Male | PDAC | I | 0.675 | 0.807 | 0.866 | 0.714 | 0.798 |
| M1165 | Female | PDAC | I | 0.849 | 0.836 | 0.887 | 0.827 | 0.878 |
| M1166 | Female | PDAC | IV | 0.773 | 0.850 | 0.674 | 0.542 | 0.661 |

| | | | | | | | | |
|---|---|---|---|---|---|---|---|---|
| M1169 | Male | PDAC | IV | 0.556 | 0.734 | 0.690 | 0.519 | 0.693 |
| M1173 | Female | PDAC | II | 0.796 | 0.917 | 0.661 | 0.293 | 0.625 |
| M1174 | Female | PDAC | III | 0.576 | 0.747 | 0.515 | 0.484 | 0.582 |
| M1175 | Male | PDAC | II | 0.440 | 0.750 | 0.430 | 0.423 | 0.542 |
| M1178 | Female | PDAC | II | 0.570 | 0.872 | 0.643 | 0.414 | 0.672 |
| M1179 | Female | PDAC | III | 0.477 | 0.873 | 0.659 | 0.476 | 0.732 |
| M1180 | Male | PDAC | IV | 0.640 | 0.774 | 0.640 | 0.316 | 0.637 |
| M1181 | Female | PDAC | I | 0.792 | 0.807 | 0.535 | 0.586 | 0.661 |
| M1183 | Male | PDAC | III | 0.678 | 0.874 | 0.788 | 0.791 | 0.842 |
| M1184 | Female | PDAC | III | 0.714 | 0.940 | 0.674 | 0.609 | 0.789 |
| M1188 | Male | PDAC | II | 0.384 | 0.930 | 0.541 | 0.736 | 0.828 |
| M1189 | Male | PDAC | III | 0.788 | 0.954 | 0.806 | 0.433 | 0.763 |
| M1191 | Male | PDAC | III | 0.594 | 0.935 | 0.796 | 0.785 | 0.892 |
| M1194 | Female | PDAC | III | 0.536 | 0.716 | 0.519 | 0.478 | 0.626 |
| M1197 | Female | PDAC | I | 0.635 | 0.878 | 0.582 | 0.363 | 0.723 |
| M1199 | Male | PDAC | II | 0.800 | 0.808 | 0.607 | 0.893 | 0.868 |
| M1200 | Female | PDAC | II | 0.316 | 0.548 | 0.381 | 0.398 | 0.502 |
| M1201 | Male | PDAC | III | 0.204 | 0.756 | 0.422 | 0.449 | 0.571 |
| M1205 | Male | PDAC | II | 0.654 | 0.771 | 0.484 | 0.265 | 0.583 |
| M1209 | Female | PDAC | III | 0.456 | 0.677 | 0.617 | 0.549 | 0.670 |
| M1214 | Female | PDAC | II | 0.591 | 0.901 | 0.718 | 0.510 | 0.768 |
| S2001 | Female | PDAC | III | 0.894 | 0.923 | 0.748 | 0.536 | 0.759 |
| S2003 | Female | PDAC | IV | 0.883 | 0.881 | 0.937 | 0.909 | 0.912 |
| S2004 | Male | PDAC | I | 0.367 | 0.719 | 0.470 | 0.589 | 0.648 |
| S2008 | Male | PDAC | III | 0.714 | 0.858 | 0.889 | 0.918 | 0.900 |
| S2011 | Male | PDAC | III | 0.832 | 0.878 | 0.549 | 0.836 | 0.837 |
| S2013 | Female | PDAC | IV | 0.640 | 0.849 | 0.487 | 0.485 | 0.677 |
| S2017 | Female | PDAC | III | 0.776 | 0.874 | 0.638 | 0.867 | 0.866 |
| S2020 | Female | PDAC | IV | 0.332 | 0.577 | 0.359 | 0.492 | 0.463 |
| S2023 | Male | PDAC | I | 0.807 | 0.796 | 0.894 | 0.904 | 0.904 |
| S2026 | Female | PDAC | IV | 0.346 | 0.458 | 0.541 | 0.579 | 0.521 |
| S2038 | Male | PDAC | IV | 0.955 | 0.937 | 0.756 | 0.687 | 0.776 |
| S2039 | Female | PDAC | IV | 0.642 | 0.723 | 0.586 | 0.531 | 0.621 |
| S2041 | Female | PDAC | II | 0.375 | 0.565 | 0.547 | 0.577 | 0.561 |
| S2045 | Female | PDAC | I | 0.638 | 0.543 | 0.503 | 0.733 | 0.638 |
| S2046 | Male | PDAC | II | 0.943 | 0.954 | 0.948 | 0.919 | 0.941 |
| S2053 | Male | PDAC | I | 0.463 | 0.376 | 0.522 | 0.397 | 0.325 |
| S2058 | Male | PDAC | IV | 0.298 | 0.492 | 0.490 | 0.611 | 0.513 |
| S2068 | Female | PDAC | III | 0.664 | 0.394 | 0.682 | 0.843 | 0.727 |

| | | | | | | | | |
|---|---|---|---|---|---|---|---|---|
| S2079 | Male | PDAC | I | 0.882 | 0.924 | 0.346 | 0.408 | 0.597 |
| S2080 | Female | PDAC | IV | 0.948 | 0.933 | 0.692 | 0.754 | 0.816 |
| S2082 | Female | PDAC | IV | 0.955 | 0.849 | 0.588 | 0.871 | 0.889 |
| S2085 | Female | PDAC | I | 0.410 | 0.448 | 0.517 | 0.591 | 0.528 |
| S2086 | Male | PDAC | II | 0.911 | 0.936 | 0.706 | 0.801 | 0.838 |
| S2088 | Male | PDAC | II | 0.614 | 0.511 | 0.632 | 0.776 | 0.640 |
| S2099 | Female | PDAC | II | 0.162 | 0.295 | 0.625 | 0.356 | 0.317 |
| S2100 | Male | PDAC | I | 0.761 | 0.747 | 0.322 | 0.649 | 0.655 |
| S2106 | Female | PDAC | IV | 0.913 | 0.839 | 0.877 | 0.911 | 0.901 |
| S2110 | Female | PDAC | II | 0.422 | 0.380 | 0.576 | 0.838 | 0.685 |
| S2116 | Female | PDAC | II | 0.883 | 0.884 | 0.705 | 0.874 | 0.866 |
| S2121 | Female | PDAC | II | 0.893 | 0.872 | 0.894 | 0.889 | 0.910 |
| S2125 | Male | PDAC | II | 0.935 | 0.911 | 0.886 | 0.881 | 0.914 |
| S2130 | Female | PDAC | I | 0.534 | 0.507 | 0.366 | 0.622 | 0.498 |
| S2132 | Male | PDAC | II | 0.638 | 0.442 | 0.648 | 0.706 | 0.618 |
| S2133 | Male | PDAC | II | 0.234 | 0.514 | 0.536 | 0.699 | 0.608 |
| S2137 | Male | PDAC | I | 0.590 | 0.381 | 0.457 | 0.676 | 0.513 |
| S2141 | Male | PDAC | III | 0.625 | 0.362 | 0.540 | 0.702 | 0.537 |
| S2144 | Male | PDAC | II | 0.954 | 0.900 | 0.606 | 0.812 | 0.823 |
| S2145 | Female | PDAC | II | 0.955 | 0.917 | 0.848 | 0.901 | 0.910 |
| S2153 | Female | PDAC | I | 0.622 | 0.720 | 0.391 | 0.744 | 0.702 |
| S2164 | Female | PDAC | II | 0.942 | 0.874 | 0.927 | 0.918 | 0.922 |
| S2165 | Male | PDAC | III | 0.821 | 0.853 | 0.537 | 0.694 | 0.734 |
| S2166 | Male | PDAC | I | 0.454 | 0.274 | 0.508 | 0.421 | 0.375 |
| S2169 | Male | PDAC | I | 0.610 | 0.813 | 0.499 | 0.457 | 0.590 |
| S2173 | Female | PDAC | II | 0.926 | 0.925 | 0.904 | 0.840 | 0.897 |
| S2175 | Male | PDAC | I | 0.233 | 0.450 | 0.332 | 0.397 | 0.361 |
| S2189 | Male | PDAC | II | 0.712 | 0.591 | 0.607 | 0.555 | 0.569 |
| S2191 | Male | PDAC | III | 0.721 | 0.573 | 0.477 | 0.629 | 0.600 |
| S2194 | Female | PDAC | I | 0.788 | 0.380 | 0.598 | 0.828 | 0.706 |
| S2195 | Male | PDAC | III | 0.955 | 0.909 | 0.650 | 0.666 | 0.776 |
| S2197 | Female | PDAC | I | 0.807 | 0.873 | 0.643 | 0.684 | 0.768 |
| S2198 | Female | PDAC | I | 0.929 | 0.925 | 0.531 | 0.570 | 0.677 |
| S2202 | Female | PDAC | II | 0.914 | 0.913 | 0.731 | 0.863 | 0.858 |
| S2209 | Male | PDAC | III | 0.432 | 0.546 | 0.445 | 0.682 | 0.606 |
| S2210 | Female | PDAC | III | 0.640 | 0.475 | 0.443 | 0.710 | 0.597 |
| S2222 | Female | PDAC | III | 0.938 | 0.815 | 0.925 | 0.892 | 0.896 |
| S2223 | Male | PDAC | I | 0.565 | 0.421 | 0.562 | 0.548 | 0.471 |
| S2225 | Female | PDAC | III | 0.820 | 0.882 | 0.917 | 0.886 | 0.917 |

| | | | | | | | | |
|---|---|---|---|---|---|---|---|---|
| S2226 | Male | PDAC | II | 0.430 | 0.485 | 0.656 | 0.855 | 0.704 |
| S2227 | Female | PDAC | II | 0.870 | 0.722 | 0.931 | 0.913 | 0.868 |
| S2228 | Female | PDAC | II | 0.968 | 0.932 | 0.710 | 0.786 | 0.847 |
| S2236 | Female | PDAC | III | 0.927 | 0.852 | 0.869 | 0.849 | 0.880 |
| S2239 | Male | PDAC | II | 0.760 | 0.291 | 0.569 | 0.805 | 0.637 |
| S2244 | Male | PDAC | I | 0.482 | 0.634 | 0.522 | 0.661 | 0.635 |
| S2247 | Female | PDAC | III | 0.746 | 0.601 | 0.602 | 0.858 | 0.791 |
| S2030 | Female | PDAC | IV | 0.786 | 0.849 | 0.622 | 0.812 | 0.830 |
| S2117 | Female | PDAC | IV | 0.971 | 0.961 | 0.691 | 0.783 | 0.837 |
| H0001 | Female | Healthy | N/A | 0.546 | 0.108 | 0.220 | 0.227 | 0.164 |
| H0002 | Female | Healthy | N/A | 0.459 | 0.321 | 0.191 | 0.333 | 0.272 |
| H0003 | Female | Healthy | N/A | 0.390 | 0.333 | 0.193 | 0.164 | 0.259 |
| H0004 | Male | Healthy | N/A | 0.637 | 0.460 | 0.295 | 0.447 | 0.347 |
| H0005 | Male | Healthy | N/A | 0.247 | 0.256 | 0.387 | 0.416 | 0.310 |
| H0007 | Male | Healthy | N/A | 0.284 | 0.174 | 0.163 | 0.317 | 0.183 |
| H0010 | Female | Healthy | N/A | 0.574 | 0.237 | 0.224 | 0.149 | 0.244 |
| H0011 | Female | Healthy | N/A | 0.507 | 0.144 | 0.349 | 0.345 | 0.214 |
| H0012 | Female | Healthy | N/A | 0.093 | 0.442 | 0.579 | 0.575 | 0.557 |
| H0015 | Male | Healthy | N/A | 0.610 | 0.442 | 0.377 | 0.254 | 0.324 |
| H0016 | Female | Healthy | N/A | 0.070 | 0.051 | 0.365 | 0.404 | 0.242 |
| H0018 | Male | Healthy | N/A | 0.319 | 0.309 | 0.375 | 0.326 | 0.246 |
| H0020 | Female | Healthy | N/A | 0.387 | 0.173 | 0.296 | 0.428 | 0.281 |
| H0021 | Female | Healthy | N/A | 0.529 | 0.261 | 0.513 | 0.246 | 0.247 |
| H0022 | Female | Healthy | N/A | 0.223 | 0.093 | 0.393 | 0.381 | 0.213 |
| H0023 | Female | Healthy | N/A | 0.684 | 0.327 | 0.461 | 0.426 | 0.292 |
| H0025 | Female | Healthy | N/A | 0.238 | 0.203 | 0.645 | 0.288 | 0.264 |
| H0026 | Male | Healthy | N/A | 0.652 | 0.416 | 0.703 | 0.265 | 0.459 |
| H0027 | Male | Healthy | N/A | 0.346 | 0.074 | 0.645 | 0.481 | 0.351 |
| H0029 | Male | Healthy | N/A | 0.303 | 0.152 | 0.395 | 0.457 | 0.317 |
| H0030 | Female | Healthy | N/A | 0.365 | 0.057 | 0.535 | 0.282 | 0.211 |
| H0031 | Female | Healthy | N/A | 0.170 | 0.052 | 0.277 | 0.328 | 0.165 |
| H0032 | Male | Healthy | N/A | 0.343 | 0.237 | 0.509 | 0.296 | 0.306 |
| H0033 | Female | Healthy | N/A | 0.246 | 0.041 | 0.353 | 0.401 | 0.228 |
| H0034 | Female | Healthy | N/A | 0.422 | 0.277 | 0.677 | 0.490 | 0.454 |
| H0036 | Female | Healthy | N/A | 0.354 | 0.105 | 0.568 | 0.532 | 0.372 |
| H0037 | Female | Healthy | N/A | 0.103 | 0.058 | 0.485 | 0.313 | 0.193 |
| H0039 | Male | Healthy | N/A | 0.428 | 0.113 | 0.298 | 0.214 | 0.177 |
| H0040 | Female | Healthy | N/A | 0.156 | 0.013 | 0.533 | 0.387 | 0.246 |
| H0041 | Female | Healthy | N/A | 0.430 | 0.185 | 0.389 | 0.548 | 0.404 |

| | | | | | | | | |
|---|---|---|---|---|---|---|---|---|
| H0042 | Male | Healthy | N/A | 0.150 | 0.050 | 0.168 | 0.429 | 0.188 |
| H0044 | Male | Healthy | N/A | 0.360 | 0.079 | 0.502 | 0.353 | 0.264 |
| H0045 | Male | Healthy | N/A | 0.257 | 0.093 | 0.518 | 0.445 | 0.286 |
| H0047 | Male | Healthy | N/A | 0.255 | 0.124 | 0.393 | 0.597 | 0.374 |
| H0048 | Male | Healthy | N/A | 0.339 | 0.547 | 0.612 | 0.467 | 0.537 |
| H0049 | Female | Healthy | N/A | 0.440 | 0.661 | 0.306 | 0.406 | 0.436 |
| H0051 | Male | Healthy | N/A | 0.603 | 0.680 | 0.212 | 0.312 | 0.469 |
| H0052 | Male | Healthy | N/A | 0.210 | 0.397 | 0.236 | 0.214 | 0.228 |
| H0053 | Female | Healthy | N/A | 0.526 | 0.372 | 0.374 | 0.309 | 0.348 |
| H0054 | Female | Healthy | N/A | 0.468 | 0.595 | 0.340 | 0.231 | 0.407 |
| H0055 | Male | Healthy | N/A | 0.758 | 0.772 | 0.484 | 0.664 | 0.725 |
| H0056 | Male | Healthy | N/A | 0.205 | 0.347 | 0.255 | 0.273 | 0.298 |
| H0058 | Female | Healthy | N/A | 0.363 | 0.115 | 0.269 | 0.306 | 0.174 |
| H0060 | Female | Healthy | N/A | 0.525 | 0.449 | 0.337 | 0.458 | 0.412 |
| H0063 | Male | Healthy | N/A | 0.232 | 0.224 | 0.310 | 0.238 | 0.214 |
| H0065 | Female | Healthy | N/A | 0.686 | 0.224 | 0.373 | 0.374 | 0.266 |
| H0066 | Female | Healthy | N/A | 0.126 | 0.037 | 0.397 | 0.349 | 0.207 |
| H0068 | Male | Healthy | N/A | 0.469 | 0.188 | 0.436 | 0.407 | 0.310 |
| H0069 | Male | Healthy | N/A | 0.197 | 0.427 | 0.356 | 0.272 | 0.336 |
| H0071 | Female | Healthy | N/A | 0.373 | 0.285 | 0.457 | 0.257 | 0.309 |
| H0073 | Female | Healthy | N/A | 0.329 | 0.207 | 0.304 | 0.162 | 0.207 |
| H0075 | Female | Healthy | N/A | 0.069 | 0.026 | 0.252 | 0.458 | 0.238 |
| H0077 | Female | Healthy | N/A | 0.141 | 0.078 | 0.304 | 0.377 | 0.203 |
| H0079 | Female | Healthy | N/A | 0.420 | 0.248 | 0.218 | 0.384 | 0.266 |
| H0081 | Female | Healthy | N/A | 0.225 | 0.148 | 0.281 | 0.305 | 0.206 |
| H0084 | Male | Healthy | N/A | 0.135 | 0.069 | 0.346 | 0.299 | 0.147 |
| H0089 | Female | Healthy | N/A | 0.509 | 0.385 | 0.235 | 0.248 | 0.283 |
| H0091 | Female | Healthy | N/A | 0.174 | 0.115 | 0.245 | 0.490 | 0.284 |
| H0092 | Male | Healthy | N/A | 0.229 | 0.369 | 0.271 | 0.208 | 0.231 |
| H0093 | Male | Healthy | N/A | 0.118 | 0.029 | 0.453 | 0.257 | 0.147 |
| H0094 | Male | Healthy | N/A | 0.238 | 0.032 | 0.462 | 0.518 | 0.297 |
| H0097 | Female | Healthy | N/A | 0.467 | 0.343 | 0.433 | 0.251 | 0.255 |
| H0099 | Female | Healthy | N/A | 0.178 | 0.193 | 0.161 | 0.158 | 0.151 |
| H0100 | Female | Healthy | N/A | 0.111 | 0.033 | 0.218 | 0.229 | 0.102 |
| H0101 | Male | Healthy | N/A | 0.099 | 0.259 | 0.200 | 0.332 | 0.231 |
| H0102 | Male | Healthy | N/A | 0.709 | 0.630 | 0.366 | 0.749 | 0.680 |
| H0103 | Male | Healthy | N/A | 0.180 | 0.221 | 0.377 | 0.439 | 0.288 |
| H0104 | Male | Healthy | N/A | 0.171 | 0.275 | 0.383 | 0.332 | 0.274 |
| H0105 | Male | Healthy | N/A | 0.119 | 0.393 | 0.243 | 0.296 | 0.242 |

| | | | | | | | | |
|---|---|---|---|---|---|---|---|---|
| H0107 | Male | Healthy | N/A | 0.521 | 0.322 | 0.298 | 0.690 | 0.527 |
| H0108 | Male | Healthy | N/A | 0.117 | 0.238 | 0.308 | 0.251 | 0.194 |
| H0109 | Female | Healthy | N/A | 0.701 | 0.672 | 0.305 | 0.256 | 0.423 |
| H0111 | Female | Healthy | N/A | 0.314 | 0.263 | 0.290 | 0.288 | 0.226 |
| H0113 | Female | Healthy | N/A | 0.110 | 0.091 | 0.175 | 0.443 | 0.221 |
| H0118 | Male | Healthy | N/A | 0.141 | 0.068 | 0.321 | 0.161 | 0.118 |
| H0119 | Female | Healthy | N/A | 0.240 | 0.184 | 0.319 | 0.527 | 0.333 |
| H0120 | Male | Healthy | N/A | 0.220 | 0.155 | 0.249 | 0.531 | 0.311 |
| H0122 | Female | Healthy | N/A | 0.493 | 0.402 | 0.198 | 0.418 | 0.369 |
| H0124 | Male | Healthy | N/A | 0.211 | 0.148 | 0.268 | 0.278 | 0.161 |
| H0126 | Female | Healthy | N/A | 0.216 | 0.212 | 0.290 | 0.254 | 0.192 |
| H0128 | Male | Healthy | N/A | 0.566 | 0.160 | 0.554 | 0.573 | 0.386 |
| H0130 | Male | Healthy | N/A | 0.178 | 0.078 | 0.259 | 0.367 | 0.212 |
| H0131 | Male | Healthy | N/A | 0.524 | 0.302 | 0.348 | 0.305 | 0.236 |
| H0134 | Female | Healthy | N/A | 0.093 | 0.016 | 0.273 | 0.430 | 0.220 |
| H0135 | Female | Healthy | N/A | 0.262 | 0.434 | 0.372 | 0.711 | 0.569 |
| H0136 | Female | Healthy | N/A | 0.233 | 0.204 | 0.379 | 0.342 | 0.224 |
| H0137 | Female | Healthy | N/A | 0.261 | 0.255 | 0.435 | 0.585 | 0.416 |
| H0138 | Male | Healthy | N/A | 0.373 | 0.142 | 0.289 | 0.538 | 0.317 |
| H0139 | Female | Healthy | N/A | 0.111 | 0.100 | 0.290 | 0.587 | 0.364 |
| H0141 | Female | Healthy | N/A | 0.271 | 0.148 | 0.332 | 0.417 | 0.266 |
| H0142 | Female | Healthy | N/A | 0.296 | 0.145 | 0.332 | 0.276 | 0.186 |
| H0143 | Female | Healthy | N/A | 0.449 | 0.702 | 0.197 | 0.152 | 0.493 |
| H0145 | Female | Healthy | N/A | 0.050 | 0.021 | 0.306 | 0.297 | 0.128 |
| H0148 | Male | Healthy | N/A | 0.089 | 0.012 | 0.325 | 0.230 | 0.095 |
| H0153 | Female | Healthy | N/A | 0.413 | 0.411 | 0.311 | 0.328 | 0.350 |
| H0154 | Male | Healthy | N/A | 0.186 | 0.058 | 0.389 | 0.237 | 0.144 |
| H0157 | Male | Healthy | N/A | 0.344 | 0.105 | 0.330 | 0.335 | 0.172 |
| H0158 | Female | Healthy | N/A | 0.766 | 0.355 | 0.586 | 0.492 | 0.481 |
| H0159 | Male | Healthy | N/A | 0.538 | 0.425 | 0.243 | 0.404 | 0.347 |
| H0165 | Male | Healthy | N/A | 0.228 | 0.172 | 0.572 | 0.365 | 0.340 |
| H0166 | Male | Healthy | N/A | 0.193 | 0.180 | 0.419 | 0.591 | 0.418 |
| H0167 | Female | Healthy | N/A | 0.078 | 0.055 | 0.423 | 0.422 | 0.228 |
| H0168 | Female | Healthy | N/A | 0.803 | 0.454 | 0.773 | 0.805 | 0.732 |
| H0174 | Female | Healthy | N/A | 0.127 | 0.095 | 0.358 | 0.351 | 0.231 |
| H0175 | Female | Healthy | N/A | 0.095 | 0.068 | 0.320 | 0.362 | 0.196 |
| H0176 | Male | Healthy | N/A | 0.423 | 0.170 | 0.433 | 0.477 | 0.361 |
| H0177 | Male | Healthy | N/A | 0.189 | 0.141 | 0.550 | 0.662 | 0.471 |
| H0178 | Female | Healthy | N/A | 0.195 | 0.066 | 0.354 | 0.293 | 0.147 |

| | | | | | | | | |
|---|---|---|---|---|---|---|---|---|
| H0179 | Female | Healthy | N/A | 0.116 | 0.033 | 0.381 | 0.331 | 0.164 |
| H0180 | Male | Healthy | N/A | 0.741 | 0.391 | 0.616 | 0.636 | 0.588 |
| H0181 | Male | Healthy | N/A | 0.149 | 0.015 | 0.430 | 0.261 | 0.141 |
| H0184 | Male | Healthy | N/A | 0.363 | 0.118 | 0.438 | 0.498 | 0.302 |
| H0188 | Female | Healthy | N/A | 0.444 | 0.617 | 0.471 | 0.295 | 0.441 |
| H0189 | Female | Healthy | N/A | 0.607 | 0.696 | 0.450 | 0.325 | 0.490 |
| H0191 | Male | Healthy | N/A | 0.765 | 0.609 | 0.314 | 0.580 | 0.591 |
| H0192 | Female | Healthy | N/A | 0.193 | 0.312 | 0.318 | 0.342 | 0.321 |
| H0198 | Female | Healthy | N/A | 0.199 | 0.436 | 0.284 | 0.383 | 0.368 |
| H0201 | Male | Healthy | N/A | 0.354 | 0.365 | 0.291 | 0.297 | 0.293 |
| H0205 | Male | Healthy | N/A | 0.223 | 0.233 | 0.468 | 0.386 | 0.265 |
| H0206 | Male | Healthy | N/A | 0.502 | 0.666 | 0.658 | 0.331 | 0.592 |
| H0209 | Male | Healthy | N/A | 0.290 | 0.367 | 0.343 | 0.344 | 0.277 |

Abbreviations: AJCC, American Joint Commission on Cancer

**Supplementary Table S4:** Protein biomarker and plasma DNA fragmentation scores for individual patients with incidental pancreas lesions.

| Patient ID | Sex | Pathology | Surgery | CA 19-9 (U/mL) | CA 125 (U/mL) | CEA (ng/ mL) | Plasma DNA fragmentation score | Multianalyte regression score |
|---|---|---|---|---|---|---|---|---|
| P2 | Female | Benign | Yes | 32.2 | 12.4 | 1.29 | 0.443 | 0.073 |
| P3 | Male | Benign | No | 11.6 | 6.7 | 1.88 | 0.42 | 0.014 |
| P5 | Female | Benign | No | 4.1 | 9 | 2.83 | 0.415 | 0.007 |
| P6 | Female | Benign | No | 5.9 | 13 | 1.23 | 0.372 | 0.012 |
| P7 | Female | Benign | No | 5.5 | 17.6 | 3.04 | 0.702 | 0.159 |
| P8 | Female | Benign | No | 21.7 | 16.2 | 2.47 | 0.466 | 0.04 |
| P9 | Male | Benign | No | 15 | 13.9 | 1.96 | 0.358 | 0.011 |
| P11 | Male | Benign | No | 6.5 | 11.8 | 1.32 | 0.495 | 0.039 |
| P13 | Female | Benign | Yes | 2.1 | 16.6 | 1.84 | 0.489 | 0.029 |
| P14 | Female | Malignant | Yes | 460 | 6.6 | 7.03 | 0.944 | 1 |
| P16 | Female | Benign | No | 2.1 | 22.4 | 1 | 0.539 | 0.085 |
| P18 | Female | Benign | No | 40.3 | 36.8 | 20.54 | 0.492 | 0 |
| P19 | Female | Malignant | Yes | 4 | 15.8 | 3.93 | 0.7 | 0.097 |
| P22 | Female | Benign | No | 10.3 | 11.1 | 1.88 | 0.676 | 0.193 |
| P23 | Female | Benign | No | 2.9 | 10.3 | 3.67 | 0.3 | 0.001 |
| P24 | Male | Benign | No | 26.4 | 10.3 | 2.8 | 0.567 | 0.092 |
| P25 | Female | Benign | Yes | 4.1 | 18.1 | 3.25 | 0.496 | 0.019 |
| P29 | Male | Benign | No | 2.1 | 51.1 | 2.79 | 0.577 | 0.16 |
| P31 | Male | Benign | No | 14 | 7.1 | 1.74 | 0.408 | 0.015 |
| P33 | Female | Benign | No | 3.2 | 17.7 | 1.69 | 0.43 | 0.018 |
| P34 | Female | Benign | No | 12.3 | 18.8 | 3.03 | 0.481 | 0.026 |
| P35 | Female | Benign | No | 32.7 | 17.3 | 4.65 | 0.512 | 0.04 |
| P36 | Female | Benign | No | 2.7 | 5.3 | 1.3 | 0.485 | 0.023 |
| P37 | Female | Benign | No | 3.6 | 26.3 | 1.05 | 0.542 | 0.106 |
| P38 | Male | Benign | No | 6 | 14.2 | 1.3 | 0.372 | 0.012 |
| P39 | Female | Benign | No | 7.6 | 22 | 1.22 | 0.419 | 0.029 |
| P41 | Male | Benign | No | 62.3 | 18.6 | 10.95 | 0.592 | 0.02 |
| P43 | Male | Benign | No | 7.9 | 10.8 | 1.19 | 0.435 | 0.023 |
| P44 | Female | Benign | No | 11.5 | 12.9 | 4.22 | 0.663 | 0.074 |
| P45 | Female | Benign | No | 38.8 | 10.4 | 8.93 | 0.497 | 0.005 |
| P47 | Male | Benign | No | 26.7 | 14.7 | 1 | 0.347 | 0.027 |
| P49 | Female | Malignant | Yes | 53.3 | 8.9 | 1 | 0.629 | 0.595 |
| P50 | Male | Benign | No | 23.1 | 31.7 | 8.55 | 0.83 | 0.176 |
| P51 | Female | Benign | No | 4 | 6.9 | 2.33 | 0.467 | 0.014 |
| P53 | Male | Benign | No | 36.4 | 9.5 | 3.48 | 0.667 | 0.242 |
| P54 | Male | Benign | No | 2.4 | 15 | 1.07 | 0.583 | 0.098 |
| P55 | Female | Benign | No | 3.7 | 45.6 | 1.66 | 0.472 | 0.085 |
| P56 | Female | Benign | No | 9.9 | 17.4 | 1.75 | 0.194 | 0.002 |
| P58 | Female | Benign | No | 19.3 | 15.4 | 2.17 | 0.511 | 0.062 |
| P59 | Female | Malignant | Yes | 39.4 | 51.3 | 2.47 | 0.456 | 0.257 |

| | | | | | | | | |
|---|---|---|---|---|---|---|---|---|
| P60 | Female | Benign | No | 14 | 8.2 | 4.06 | 0.438 | 0.007 |
| P61 | Male | Benign | No | 5.8 | 5.5 | 1.69 | 0.367 | 0.007 |
| P62 | Female | Benign | No | 2.1 | 21.9 | 4.86 | 0.578 | 0.022 |
| P63 | Male | Benign | No | 27 | 13.7 | 1 | 0.732 | 0.61 |
| P64 | Female | Benign | No | 2.1 | 30 | 3.32 | 0.606 | 0.079 |
| P65 | Male | Benign | No | 9.8 | 26.2 | 1.2 | 0.485 | 0.074 |
| P66 | Male | Benign | No | 7.5 | 13.2 | 1.71 | 0.306 | 0.005 |
| P67 | Male | Benign | Yes | 3.1 | 18.3 | 1.26 | 0.542 | 0.07 |
| P68 | Male | Benign | Yes | 10.8 | 18.3 | 2.09 | 0.548 | 0.072 |
| P70 | Male | Malignant | Yes | 9 | 13.5 | 3.28 | 0.89 | 0.549 |
| P71 | Female | Benign | No | 5.3 | 11.2 | 1 | 0.341 | 0.009 |
| P73 | Male | Benign | No | 5.4 | 19.3 | 3.29 | 0.599 | 0.057 |
| P75 | Male | Benign | No | 4.4 | 9.2 | 1.9 | 0.474 | 0.02 |
| P76 | Female | Malignant | No | 1293.9 | 33 | 1.88 | 0.64 | 1 |
| P77 | Female | Benign | No | 2.1 | 26 | 3.56 | 0.313 | 0.003 |
| P78 | Female | Benign | No | 2.7 | 8.5 | 4.1 | 0.509 | 0.009 |
| P80 | Female | Benign | Yes | 2.4 | 11.9 | 1 | 0.392 | 0.013 |
| P81 | Female | Benign | No | 2.1 | 25 | 3.56 | 0.572 | 0.042 |
| P82 | Male | Benign | No | 5.9 | 18.9 | 1.83 | 0.553 | 0.07 |
| P83 | Female | Malignant | Yes | 2.1 | 21.2 | 2.08 | 0.394 | 0.012 |
| P85 | Female | Malignant | No | 15.8 | 17.3 | 1.75 | 0.55 | 0.101 |
| P86 | Male | Benign | Yes | 2.1 | 8.6 | 2.52 | 0.567 | 0.034 |
| P87 | Male | Benign | No | 47.6 | 4.7 | 1.02 | 0.315 | 0.034 |
| P88 | Female | Benign | No | 3.5 | 7.8 | 1.54 | 0.384 | 0.008 |
| P89 | Male | Benign | No | 5.7 | 16.9 | 3.66 | 0.587 | 0.04 |
| P90 | Female | Malignant | No | 1409.3 | 53.8 | 3.62 | 0.172 | 1 |
| P91 | Female | Benign | No | 7 | 17 | 1.14 | 0.232 | 0.004 |
| P92 | Male | Benign | No | 13 | 22.1 | 5.35 | 0.236 | 0.001 |
| P93 | Female | Benign | Yes | 15.7 | 10.8 | 1 | 0.373 | 0.019 |
| P94 | Female | Benign | No | 13.3 | 13.7 | 1.27 | 0.482 | 0.05 |
| P96 | Female | Benign | No | 32.7 | 15.9 | 1.15 | 0.153 | 0.005 |
| P97 | Female | Benign | No | 5 | 18.3 | 2.68 | 0.61 | 0.079 |
| P99 | Male | Benign | No | 12.9 | 7.5 | 2.24 | 0.389 | 0.01 |
| P100 | Female | Benign | No | 50.3 | 21.3 | 1.07 | 0.342 | 0.091 |
| P102 | Female | Benign | No | 2.1 | 13 | 1 | 0.573 | 0.084 |
| P103 | Male | Benign | No | 12.7 | 11 | 5.93 | 0.414 | 0.003 |
| P104 | Female | Malignant | No | 18917 | 10.2 | 3.11 | 0.571 | 1 |
| P107 | Male | Benign | Yes | 8.9 | 14.5 | 1 | 0.407 | 0.023 |
| P119 | Female | Benign | Yes | 2.9 | 5.6 | 3.46 | 0.408 | 0.004 |
| P120 | Female | Benign | No | 23.9 | 18.3 | 1.72 | 0.544 | 0.139 |
| P241 | Female | Benign | No | 12.5 | 10.9 | 1.31 | 0.336 | 0.01 |
| P242 | Female | Benign | No | 26.3 | 98.1 | 8.92 | 0.484 | 0.074 |
| P245 | Female | Benign | Yes | 2.1 | 13.4 | 1.65 | 0.379 | 0.009 |
| P247 | Female | Benign | No | 2.9 | 38.4 | 2.19 | 0.298 | 0.008 |

| | | | | | | | | |
|---|---|---|---|---|---|---|---|---|
| P249 | Female | Benign | No | 2.1 | 23 | 1 | 0.221 | 0.003 |
| P251 | Female | Benign | Yes | 8.6 | 13.2 | 1.03 | 0.575 | 0.111 |
| P253 | Female | Benign | Yes | 2.1 | 8.2 | 2.28 | 0.393 | 0.006 |
| P255 | Male | Benign | Yes | 10.1 | 87.1 | 1.51 | 0.368 | 0.191 |
| P256 | Male | Malignant | Yes | 526.1 | 9.7 | 5.25 | 0.272 | 1 |
| P257 | Male | Benign | No | 3.9 | 13.3 | 1.56 | 0.272 | 0.003 |
| P258 | Female | Benign | No | 31.7 | 21.2 | 4.81 | 0.451 | 0.022 |
| P259 | Male | Benign | No | 2.9 | 8.4 | 6.91 | 0.388 | 0.001 |
| P261 | Male | Benign | No | 72.1 | 10.2 | 5.98 | 0.513 | 0.094 |
| P262 | Male | Benign | No | 8.1 | 21.3 | 1.81 | 0.199 | 0.002 |
| P263 | Female | Benign | No | 4.3 | 15.5 | 1.84 | 0.231 | 0.002 |
| P265 | Male | Benign | No | 15.7 | 12.2 | 2.21 | 0.191 | 0.002 |
| P266 | Male | Benign | No | 12 | 13.8 | 3.82 | 0.501 | 0.018 |
| P267 | Male | Benign | No | 24 | 10.7 | 2.55 | 0.188 | 0.002 |
| P268 | Male | Benign | No | 17.9 | 8.3 | 3.31 | 0.209 | 0.001 |

**Supplementary Table S5:** Clinical information for 25 patients diagnosed with histologically confirmed pathology. Patients are ranked in descending order by multianalyte regression score.

| Patient ID | Sex | Pathology | Disease Stage | Cyst-associated | CA 19-9 (U/mL) | CA 125 (U/mL) | CEA (ng/mL) | Plasma DNA fragmentation score | Multianalyte regression score |
|---|---|---|---|---|---|---|---|---|---|
| P14 | Female | Malignant | pIIB | Yes | 460 | 6.6 | 7.03 | 0.944 | 1 |
| P256 | Male | Malignant | pIIB | Yes | 526.1 | 9.7 | 5.25 | 0.272 | 1 |
| P76 | Female | Malignant | cIIB | Yes | 1293.9 | 33 | 1.88 | 0.64 | 1 |
| P90 | Female | Malignant | cIIB | Yes | 1409.3 | 53.8 | 3.62 | 0.172 | 1 |
| P104 | Female | Malignant | cIB | Yes | 18917 | 10.2 | 3.11 | 0.571 | 1 |
| P49 | Female | Malignant | pIB | Yes | 53.3 | 8.9 | 1 | 0.629 | 0.595 |
| P70 | Male | Malignant | HGD | Yes | 9 | 13.5 | 3.28 | 0.89 | 0.549 |
| P59 | Female | Malignant | pIIA (SPT) | Yes | 39.4 | 51.3 | 2.47 | 0.456 | 0.257 |
| P255 | Male | Benign | N/A | N/A | 10.1 | 87.1 | 1.51 | 0.368 | 0.191 |
| P251 | Female | Benign | N/A | N/A | 8.6 | 13.2 | 1.03 | 0.575 | 0.111 |
| P85 | Female | Malignant | cIA | No | 15.8 | 17.3 | 1.75 | 0.55 | 0.101 |
| P19 | Female | Malignant | pIIB | Yes | 4 | 15.8 | 3.93 | 0.7 | 0.097 |
| P2 | Female | Benign | N/A | N/A | 32.2 | 12.4 | 1.29 | 0.443 | 0.073 |
| P68 | Male | Benign | N/A | N/A | 10.8 | 18.3 | 2.09 | 0.548 | 0.072 |
| P67 | Male | Benign | N/A | N/A | 3.1 | 18.3 | 1.26 | 0.542 | 0.07 |
| P86 | Male | Benign | N/A | N/A | 2.1 | 8.6 | 2.52 | 0.567 | 0.034 |
| P13 | Female | Benign | N/A | N/A | 2.1 | 16.6 | 1.84 | 0.489 | 0.029 |
| P107 | Male | Benign | N/A | N/A | 8.9 | 14.5 | 1 | 0.407 | 0.023 |
| P25 | Female | Benign | N/A | N/A | 4.1 | 18.1 | 3.25 | 0.496 | 0.019 |
| P93 | Female | Benign | N/A | N/A | 15.7 | 10.8 | 1 | 0.373 | 0.019 |
| P80 | Female | Benign | N/A | N/A | 2.4 | 11.9 | 1 | 0.392 | 0.013 |

| | | | | | | | | | |
|---|---|---|---|---|---|---|---|---|---|
| P83 | Female | Malignant | HGD | Yes | 2.1 | 21.2 | 2.08 | 0.394 | 0.012 |
| P245 | Female | Benign | N/A | N/A | 2.1 | 13.4 | 1.65 | 0.379 | 0.009 |
| P253 | Female | Benign | N/A | N/A | 2.1 | 8.2 | 2.28 | 0.393 | 0.006 |
| P119 | Female | Benign | N/A | N/A | 2.9 | 5.6 | 3.46 | 0.408 | 0.004 |

Abbreviations: HGD, high-grade dysplasia; SPT, solid pseudopapillary tumor.
